# Immuno-proteomic profiling reveals abundant airway CD8 T cells and ongoing epithelial injury in prolonged post-COVID19 respiratory disease

**DOI:** 10.1101/2021.08.10.21261834

**Authors:** Bavithra Vijayakumar, Karim Boustani, Patricia P. Ogger, Artermis Papadaki, James Tonkin, Christopher M. Orton, Poonam Ghai, Kornelija Suveizdyte, Richard J. Hewitt, Robert J. Snelgrove, Philip L. Molyneaux, Justin L. Garner, James E. Peters, Pallav L. Shah, Clare M. Lloyd, James A. Harker

**Affiliations:** National Heart and Lung Institute, Imperial College London, London, UK; Chelsea and Westminster Hospital, London, UK; Royal Brompton and Harefield Hospitals, Guy’s and St Thomas’ NHS Foundation Trust, London, UK; Asthma UK Centre for Allergic Mechanisms of Asthma; Centre for Inflammatory Disease, Dept of Immunology and Inflammation, Imperial College London, Hammersmith Hospital, London, UK

**Keywords:** respiratory viral infection, tissue resident memory, COVID19, SARS-CoV-2, airways

## Abstract

Some patients hospitalized with acute COVID19 suffer respiratory symptoms that persist for many months. To characterize the local and systemic immune responses associated with this form of ‘Long COVID’, we delineated the immune and proteomic landscape in the airway and peripheral blood of normal volunteers and patients from 3 to 6 months after hospital discharge. The bronchoalveolar lavage (but not peripheral blood) proteome was abnormal in patients with post-COVID19 lung disease with significantly elevated concentration of proteins associated with apoptosis, tissue repair and epithelial injury. This correlated with an increase in cytotoxic lymphocytes (especially tissue resident CD8^+^ T cells), lactate dehydrogenase and albumin (biomarkers of cell death and barrier integrity). Follow-up of a subset of these patients greater than 1-year post-COVID19 indicated these abnormalities resolved over time. Collectively, these data indicate that COVID-19 results in a prolonged change to the airway immune landscape in those with persistent lung disease, with evidence of cell death and tissue repair linked to ongoing activation of cytotoxic T cells.

**Highlights:**

- The post-COVID19 airway is characterized by increased cytotoxic lymphocytes.
- Distinct airway proteomes are associated with the airway immune cell landscape.
- The peripheral blood does not predict immune-proteome alterations in the airway post-COVID19.
- Persistent abnormalities in the airway immune-proteome post-COVID19 airways correlate with ongoing epithelial damage.

## Introduction

Severe acute respiratory syndrome coronavirus 2 (SARS-CoV-2) related coronavirus disease (COVID19) manifests as a spectrum of acute illnesses ranging from mild respiratory symptoms to severe, sometimes fatal, respiratory failure (Docherty et al., 2020). While the acute impact of COVID19 on morbidity and mortality is well-documented, we are still in the infancy of understanding the longer-term consequences. Morbidity from a range of persistent symptoms, including breathlessness, fatigue and memory impairment have been noted in patients recovering after the acute illness and described under the umbrella term of “long COVID” (Nalbandian et al., 2021; Sigfrid et al., 2021). Complex respiratory complications have been found in up to 18.4% of inpatients (Drake et al., 2021), and persistent breathlessness reported in more than 50% of patients recovering from COVID19 (Mandal et al., 2021). The underlying aetiology for persistent respiratory morbidity is likely to be multifactorial but may be due to persistent parenchymal abnormalities and resultant ineffective gaseous exchange. Persistent radiological abnormalities post-COVID19 are common and may be present even up to 6 months post hospital discharge (Fabbri et al., 2021; Guler et al., 2021; Han et al., 2021; Myall et al., 2021). There is, therefore, a pressing need to understand the molecular and cellular basis of post-COVID19 pulmonary syndromes.

The acute immunological and inflammatory events that occur during human respiratory virus infections, including SARS-CoV-2, are relatively well described (Harker and Lloyd, 2021). In contrast, the immunological landscape of the human respiratory tract after recovery from acute viral infection is poorly understood. SARS-CoV-2 infection results in formation of long-lasting systemic immunological memory, with virus-specific antibodies and T cell responses still detectable in the majority of those infected at least 8 months post infection and higher titers seen in previously hospitalized individuals (Dan et al., 2021). Circulating lymphocyte counts and the function and frequency of monocytes are also reduced during acute disease, but they appear to return to normal shortly after resolution of acute disease (Mann et al., 2020; Scott et al., 2020). Likewise, plasma concentrations of inflammatory mediators such as IL-6 and CXCL10, that are highly elevated in acute disease, reduce as individuals recover (Rodriguez et al., 2020). Together, this suggests that systemic inflammatory and immune responses associated with acute disease severity resolve in line with recovery from the acute symptoms. It therefore remains unclear if the severity of inflammation during acute disease is associated with the persistent respiratory pathology seen in some SARS-CoV-2 infected individuals months after infection, or if there is ongoing inflammation in these individuals.

This study sought to examine the relationship between the immune system and respiratory pathology post-COVID19. The immune cell and proteomic composition of the airways and peripheral blood were analyzed in a group of previously hospitalized COVID19 patients with persistent radiological abnormalities in their lungs over 3 months post discharge. In comparison to healthy individuals, the post-COVID19 airway showed substantial increases in activated tissue resident memory CD8^+^ and CD4^+^ TRM, and an altered monocyte pool. The airway proteome was also distinct from that observed in healthy individuals, with elevation in proteins associated with ongoing cell death, loss of barrier integrity and immune cell recruitment. Importantly, none of these alterations were predicted by changes in the proteome or immune cells of the matched peripheral blood. Moreover, the scale of these alterations was not linked to the initial severity of disease while in hospital but instead was heterogenous. Some individuals displaying heightened T cell responses associated with significant increases in CXCR3 chemokines in the airways and linked to prolonged epithelial damage and extracellular matrix (ECM) dysregulation, while other individuals exhibited a return to relative airway homeostasis. Subsequent long-term follow-up also suggested that these ongoing changes to the airway landscape progressively return to normal.

## Results

### Increased airway lymphocyte numbers characterize patients recovering from hospitalization with SARS-CoV-2

We recruited 38 patients undergoing bronchoscopy for investigation of persistent respiratory abnormalities 3-6 months following infection after acute SARS-CoV-2 infection (post-COVID19) (**Figure 1A)**. All patients had both ongoing respiratory symptoms and radiological pulmonary abnormalities on computed tomography (CT) scanning. We obtained plasma samples from peripheral blood and bronchoalveolar lavage (BAL) fluid from their airways. We stratified the post-COVID19 cohort based on the level of respiratory support used during their initial hospitalization with acute COVID19, into moderate (no or minimal oxygen administered), severe (non-invasive ventilation) and very severe (invasive ventilation). To provide a control group, we used BAL and plasma samples obtained from 10 healthy volunteers recruited prior to the COVID19 pandemic (demographic information in **Table S1**).

**Figure 1:**
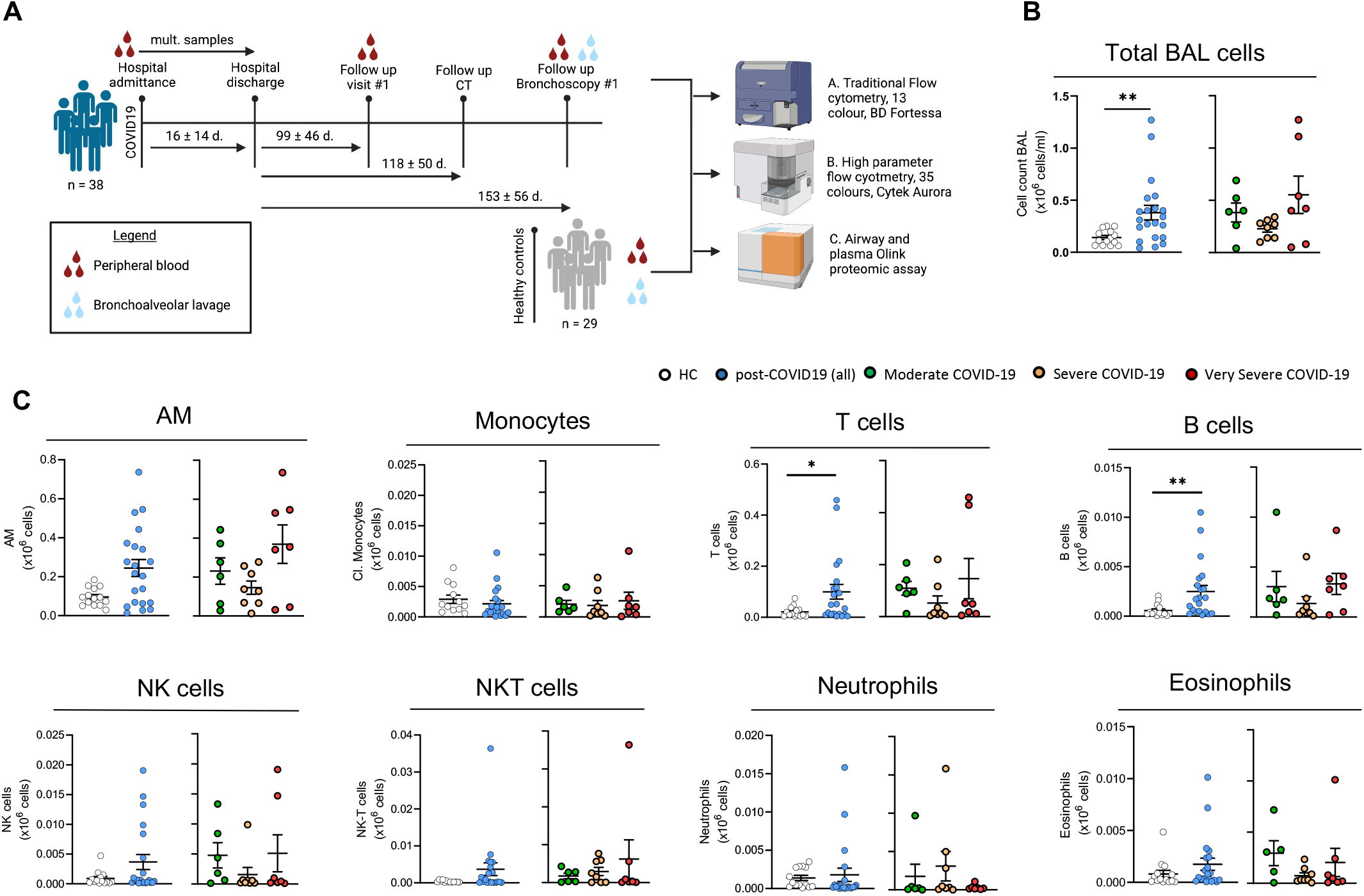
Altered immune cell profile in post-COVID19 BAL over 80 days post discharge. **(A)** Schematic of study design. BAL and blood were sampled from healthy donors and at > 80 days after hospital discharge from post-COVID19 patients. Traditional and spectral flow cytometry and Olink proteomics was performed on BAL and plasma and correlated with clinical parameters. **(B)** Left: Total number of cells in BAL from healthy controls and post-COVID19 patients. Right: total number of cells in BAL from post-COVID19 patients, stratified according to severity of the acute illness. **(C)** Total cell numbers of immune populations (x10^6^/ml) in BAL from healthy controls and post-COVID19 patients, based on gating shown in Methods Figure 1. Data are presented as mean ± SEM. Healthy controls n = 16, post-COVID19 patients n = 21, moderate n = 6, severe n = 8, very severe n = 7. Statistical significance was tested by Mann Whitney U test or Kruskal Wallis test + Dunn’s multiple comparison test. *P < 0.05, **P < 0.01, ***P < 0.005, ****P < 0.001.

We first compared the cellular composition of BAL fluid in post-COVID19 patients to healthy controls (HC) by flow cytometry. Post-COVID19 patients had a significantly higher numbers of cells in their airways compared to the healthy volunteers (**Figure 1B**). This increased cellularity was due to significantly elevated numbers of airway T and B cells, and a trend towards increased airway macrophages (AM), CD56^+^CD3^-^ (natural killer, NK) cell and CD56^+^CD3^+^ (NKT) cell numbers, while CD14^+^ monocyte, eosinophil and neutrophil numbers were similar to those found in healthy controls (**Figure 1C**). As a proportion of airway leukocytes, CD14^+^ monocytes and neutrophils were decreased in patients recovering from COVID19 compared to healthy controls (**Figure S1A**). There was no association between the severity of acute COVID19 in hospital and the immune cell composition of the post-COVID19 airways (**Figure 1C**). In contrast to the peripheral lymphopenia that is associated with acute COVID19 (Chen and Wherry, 2020), we found that in this post-COVID19 patient cohort the frequency of T cells, B cells and CD14^+^ monocytes in the peripheral blood was broadly similar to healthy controls (**Figure S1B**), although the proportion of NK and NKT cells was significantly decreased (**Figure S1B**). Collectively, these data indicate that after recovery from severe SARS-CoV-2 infection, immune cell frequencies in the peripheral blood are comparable to those in a group of age-matched controls. In contrast, the immune landscape of the airways remains altered, being marked by residual lymphocytes.

### The post-COVID19 airway immune landscape displays a limited relationship to inflammatory biomarkers found during acute disease

Severe COVID19 has a distinctive clinical biomarker blood profile, characterized by elevated inflammatory markers (including C-reactive protein (CRP) and ferritin), elevated coagulation markers (D-dimer and fibrinogen), and lymphopenia (Guan et al., 2020). At the time of hospitalization, our patients displayed a similar pattern of clinical laboratory test abnormalities (**Table S1)**. We therefore examined whether there was any association between the peak levels (or nadir in the case of lymphocyte count) of these clinical biomarkers measured during acute hospitalization and the immune cell composition of the airways at 3 months following discharge. Of the 5 biomarkers measured, ferritin correlated (Spearman rho > 0.4) with total airway monocytes, specifically classical and intermediate monocytes, and eosinophils, and inversely correlated with AM (**Table S2**). Total airway B cells correlated with peak D-dimer, while airway eosinophils correlated with lowest lymphocytes. None of the biomarkers correlated with follow up airway T cell, NK cell or neutrophil numbers, and fibrinogen and WCC showed no substantial correlation with any airway immune cell analyzed. Taken together these data suggest that the post-COVID19 airway immune cell composition shows a limited relationship with the severity of initial acute disease. This observation is supported by the lack of differences in immune cell composition when segregating between moderate, severe and very severe patients (**Figure 1**).

### The post-COVID19 airway displays a proteome profile distinct from that found in the circulation

Since clinical biomarkers did not reveal a relationship between acute disease and prolonged pathology and immune responses in the lungs post discharge, we next evaluated the airway and blood (plasma) proteomes at follow-up. We used the Olink proteomics platform to measure 435 unique proteins (**Supplementary File 1A**) in 19 post-COVID19 patients and 9 healthy volunteers. The proteins measured were highly enriched for those involved in immuno-inflammatory processes (**Supplementary File 1B-C**).

Principal components analysis (PCA) of BAL fluid proteomes revealed differences between post-COVID19 patient and healthy control samples (**Figure 2A**), with separation of case and controls most evident along PC1. In plasma, PCA also revealed differences in post-COVID19 patients and healthy controls, most evident along PC2, although the differences appeared less marked than for BAL. However, in both BAL and plasma there was considerable overlap in the spatial location of post-COVID19 and healthy control samples in the PCA plots. Unsupervised hierarchical clustering revealed two major clusters of samples in BAL, one consisting predominantly of post-COVID19 samples and the other predominantly healthy controls (**Figure S2A**), although, as with the PCA, some patient and control samples were grouped together. In contrast, in plasma, there was no visible structure to the clustering and lack of clear separation of cases and control samples (**Figure S2B**). These analyses indicate that post-COVID19 syndromes are reflected to a much greater extent in the airway proteome than the peripheral blood. They also suggest that there is considerable heterogeneity in the BAL and blood proteomes of post-COVID19 patients, with some patients displaying similar profiles to that of healthy controls, despite persistent symptoms and radiological findings.

**Figure 2:**
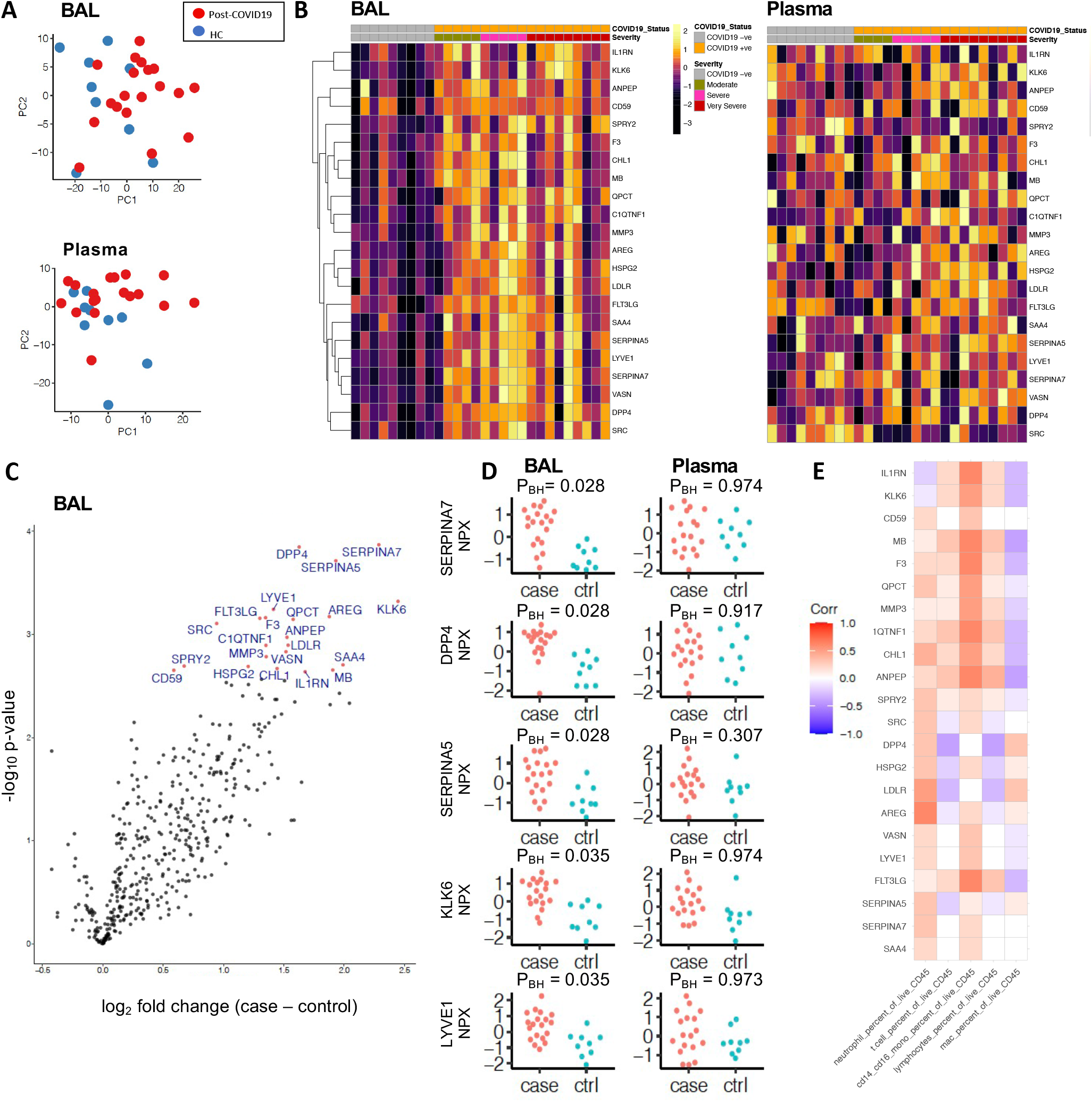
A distinct proteome is present in the post COVID-19 airway. 436 proteins in BAL and plasma 435 proteins were measured using Olink immunoassays in post-COVID19 patients (n = 19) and healthy controls (n = 9). **(A)** Principal component analysis (PCA) of BAL and plasma proteomes: each point represents a sample. **(B)** Left: heatmap displaying Z-score normalised protein abundance for the 22 proteins that were significantly differentially abundant (5% FDR) between post-COVID19 and healthy controls in BAL. Samples have been ordered by case control status and then by peak severity during acute COVID-19 infection. Proteins are ordered by hierarchical clustering. Right: heatmap for these same 22 proteins in plasma, presented in the same order as for BAL. **(C)** Volcano plot showing differentially protein abundance analysis between post-COVID19 patients and healthy controls in BAL. Nominal –log_10_ P values are shown. Significantly differentially abundant proteins (5% FDR) are coloured in red and labelled. **(D)** BAL and plasma normalised protein abundance (NPX) expression for the 5 most significantly differentially abundance proteins between post-COVID19 patients and healthy controls. P_BH_ = Benjamini-Hochberg adjusted p-values. **(E)** Correlation between the 22 differentially abundant proteins and immune cell frequency in BAL.

We next performed differential protein abundance analysis comparing post-COVID19 cases with healthy controls. In BAL fluid, we identified 22 proteins with significantly altered concentration (5% false discovery rate, FDR) (**Figure 2B-C, Supplemental File 1D**). These were all upregulated in post-COVID19 patients compared to healthy controls (**Figure 2C**). In order to provide a succinct and standardised nomenclature, we report proteins by the symbols of the genes encoding them (see **Supplementary File 1A** for a mapping of symbols to full protein names). The proteins that were most significantly differentially abundant between post-COVID19 patients and healthy controls were: SERPINA7 (thyroxine binding globulin), DPP4 (dipeptidyl peptidase 4), SERPINA5 (plasma serine protease inhibitor), KLK6 (kallikrein related peptidase-6), LYVE1 (Lymphatic vessel endothelial hyaluronic acid receptor 1), AREG (amphiregulin), F3 (factor 3), FLT3LG (Fms-related tyrosine kinase 3 ligand), QPCT (glutaminyl-peptide cyclotransferase), and SRC (Proto-oncogene tyrosine-protein kinase Src) (**Figure 2C-D**). Pathway annotation of the 22 upregulated proteins using String-DB highlighted “leucocyte activation”, “regulation of cell death”, “response to injury” and “response to wounding” (**Supplemental File 1E)**. Next, we analyzed the relationship between the 22 differentially abundant proteins and the airway immune cell proportions. Neutrophils most strongly correlated with AREG and LDLR (low density lipoprotein receptor), while monocyte proportions correlated with F3, FLT3LG, MB (myoglobin) and IL1RN (IL-1 receptor antagonist protein) (**Figure 2E)**. T cells, despite being significantly elevated in the airways of post-COVID19 patients, displayed only weak correlations with the differentially abundant proteins.

In contrast to BAL, no significant differences between protein levels were detected in plasma in post-COVID19 patients versus healthy controls (**Supplemental File 1F**). Comparison of the estimated log_2_ fold changes for the 22 proteins upregulated in post-COVID19 BAL fluid with the estimated log_2_ fold changes for these same proteins in plasma revealed no correlation (**Figure S2C-D**), indicating an absence of any relationship between airway and plasma proteome changes.

The modest sample size and multiple testing burden of 435 proteins likely limited the statistical power to detect differentially abundant proteins. To examine whether there was evidence of signal in the proteomic data that was hidden by the hard-thresholding in the differential abundance analysis, we examined quantile-quantile (QQ) plots of the distribution of expected p-values under the null hypothesis of no proteomic differences between cases and controls versus the observed p-values. For both BAL and plasma, the QQ plots revealed substantial deviation from the diagonal (albeit more so in BAL), indicating the presence of systematic differences between post-COVID19 and healthy controls for plasma proteins as well as BAL proteins (**Figure S3A)**. Corroborating this, the distribution of p-values for the proteins was not uniformly distributed, with skewing towards zero (**Figure S3B**). This is consistent with the observation that there was separation of post-COVID19 and control samples on the PCA plots for both BAL and plasma. In summary, these data suggests that there are differences in both the BAL and plasma proteomes of post-COVID19 cases compared to healthy controls, but that the effects are much stronger in BAL and this study was underpowered to detect them in plasma.

To increase power, and investigate potential protein-protein relationships, we utilized a network analysis method, Weighted Coexpression Network Analysis (WGCNA) (Langfelder and Horvath, 2008; Zhang and Horvath, 2005), that leverages the correlation between proteins to enable dimensionality reduction and thus reduce multiple testing burden. We used WGCNA to identify modules of correlated proteins, and then tested for association between these protein modules (represented quantitatively by an eigenprotein value) and case/control status. In BAL fluid, this revealed two modules (‘red’ and ‘blue’) associated with case/control status using a 5% FDR significance cut-off (**Figure 3A, Supplementary File 1G-I**). Applying a more conservative Bonferroni corrected p-value threshold, only the red module remained significant (P_Bonferonni_ 0.03).

**Figure 3:**
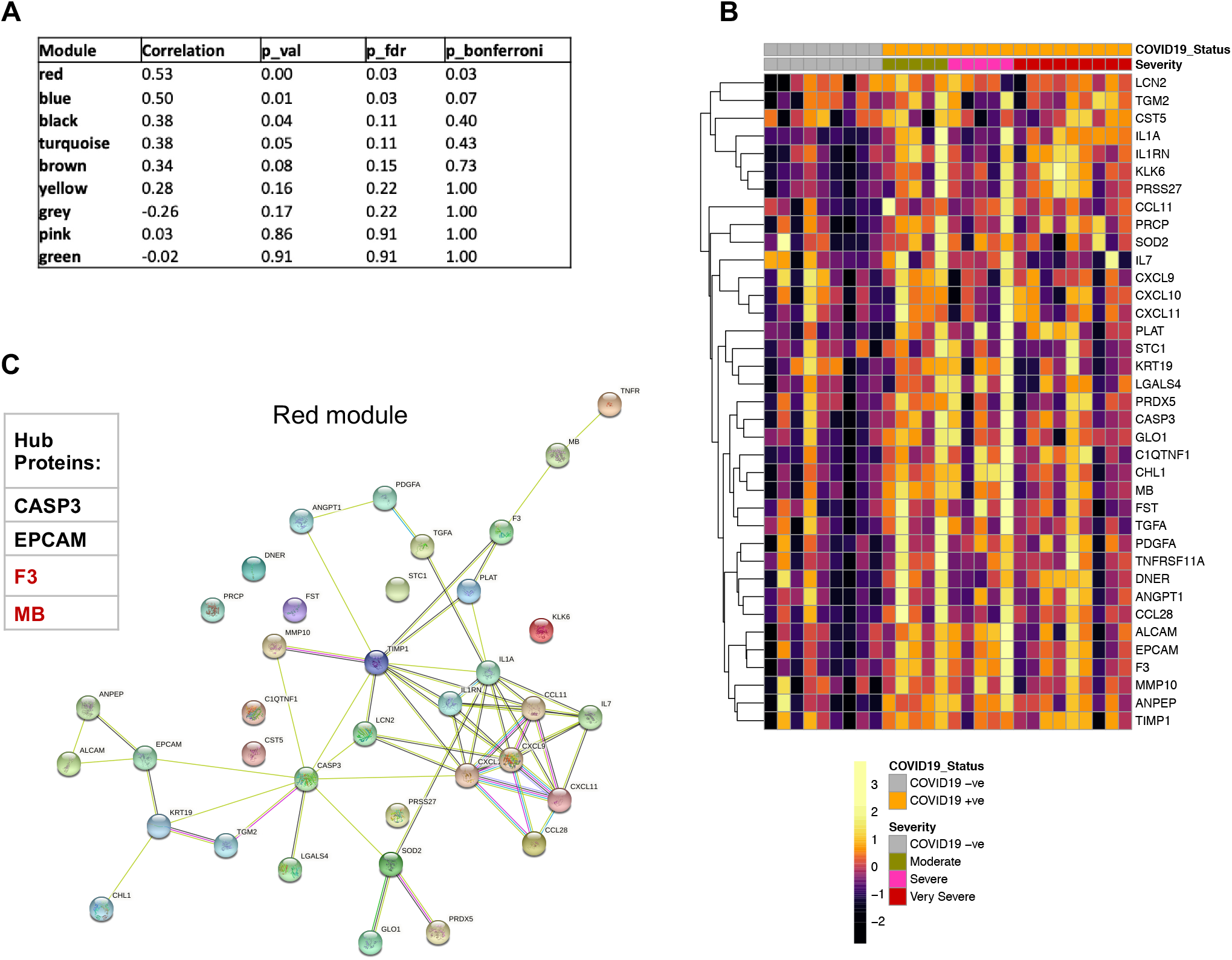
A network of proteins linked to immune cell chemotaxis and cell death is upregulated in the BAL post-COVID19. WGCNA of BAL proteome of post-COVID19 patients and healthy controls. **(A)** Associations of protein modules identified by WGCNA with case-control status (post-COVID19 or healthy). **(B)** Heatmap displaying Z-score normalised protein abundance for the 37 proteins that form the ‘red’ protein module. Samples are ordered according to clinical status. Severity refers to peak severity of the acute COVID19 episode. Proteins are ordered by hierarchical clustering. **(C)** Right: Network representation of proteins in the red module and their interconnections defined using String-db. An edge in the network represents a relationship between proteins, coloured according to the type of evidence for the connection (see Methods). Left: list of hub proteins within the network (with hub proteins that were also significantly differentially upregulated in post-COVID19 patient BAL highlighted in red).

The red module consisted of 37 proteins (**Figure 3B, Supplementary File 1H**). Overall, the red module was characterized by proteins associated with chemotaxis, inflammation, cell death and repair. Examination of the levels of proteins in the red module across samples highlighted both upregulation of individual proteins in post-COVID19 patients versus healthy controls, and the co-upregulation of groups of related proteins such as the CXCR3 chemokines (CXCL9, CXCL10 and CXCL11), and IL1A (interleukin-1A) and its antagonist IL1RN (**Figure 3B**). We used the STRING database to visualize known or predicted relationships between proteins in the module (**Figure 3C**, Methods). To highlight putative key proteins in the red and blue modules in a data-driven way, we identified hub proteins, defined as those that are highly interconnected in the proteomic network defined by WGCNA (**Supplementary File 1J**). This analysis identified CASP3 (caspase-3), EPCAM (epithelial cell adhesion molecule), F3 and MB as hub proteins in the red module. Notably, F3 and MB, an oxygen binding protein release of which is linked to muscle damage, were also identified as upregulated in the univariate differential abundance analysis (**Figure 2B-C**). CASP3 is a protein involved in cell death, EPCAM and KRT19 (Keratin-19) are indicative of epithelial cell debris within the BAL, and TGFA (transforming growth factor A) is an EGFR ligand involved in EC repair. The presence of CASP3, EPCAM, KRT19 and TGFA in the red eigenprotein module therefore suggests that one of the key features of the post-COVID19 airway is the presence of ongoing epithelial injury and repair.

As with the red module, blue module proteins were predominantly upregulated in post-COVID19 versus healthy control BAL (**Figure S4A**). The blue module was larger than the red module, containing 108 proteins involved in a wide range of biological activities. Several members were involved in cell adhesion and immune cell signaling. The hub proteins in the blue module were CD93 (Complement component C1q receptor), COMP (Cartilage oligomeric matrix protein), IGFBP3 (Insulin-like growth factor-binding protein 3), IL1R2 (Interleukin-1 receptor type 2), LYVE1, MMP2 (72 kDa type IV collagenase), NCAM1 (Neural cell adhesion molecule 1), SELL (L-selectin), TIE1 (Tyrosine-protein kinase receptor Tie-1), TNXB (Tenascin-X) and VASN (Vasorin) (**Figure S4B**). Of these, LYVE1 and VASN were also identified in the differential abundance analysis.

In contrast to the BAL network analysis, no protein modules in plasma were associated with case-control status. These results suggest that persistent post-COVID19 respiratory abnormalities have a demonstrable proteomic signature in BAL that is distinct compared to that of healthy control BAL. In contrast, we were unable to detect changes in the plasma proteome of post-COVID19 patients, even with the enhanced statistical power provided by the WGCNA method.

### CXCR3 ligands and signs of ongoing epithelial damage correlate with airway T cell and monocyte responses

Given that the airways of post-COVID19 patients displayed a distinct proteome alongside elevated numbers of T cells, B cells and NK cells, we next sought to determine which specific proteins might be linked to distinct immune cell populations. Testing for associations between BAL fluid proteins and immune cell composition in post-COVID19 patients revealed several significant findings (**Figure 4A and Supplemental file 1K**). The proportion of monocytes in the airways was significantly associated with a range of airway proteins, including the CCR7 ligand CCL19, the CXCR3 ligands CXCL9 and 11, TRAIL (TNFSF10), and BAFF (TNFSF13B) (**Figure 4A**). CXCL9 and 11 also positively correlated with lymphocyte and T cell frequencies and negatively correlated with airway macrophage frequencies in the BAL (**Figure 4A**). T cell frequencies were also positively correlated with SH2D1A (otherwise known as SLAM associated protein or SAP).

**Figure 4.**
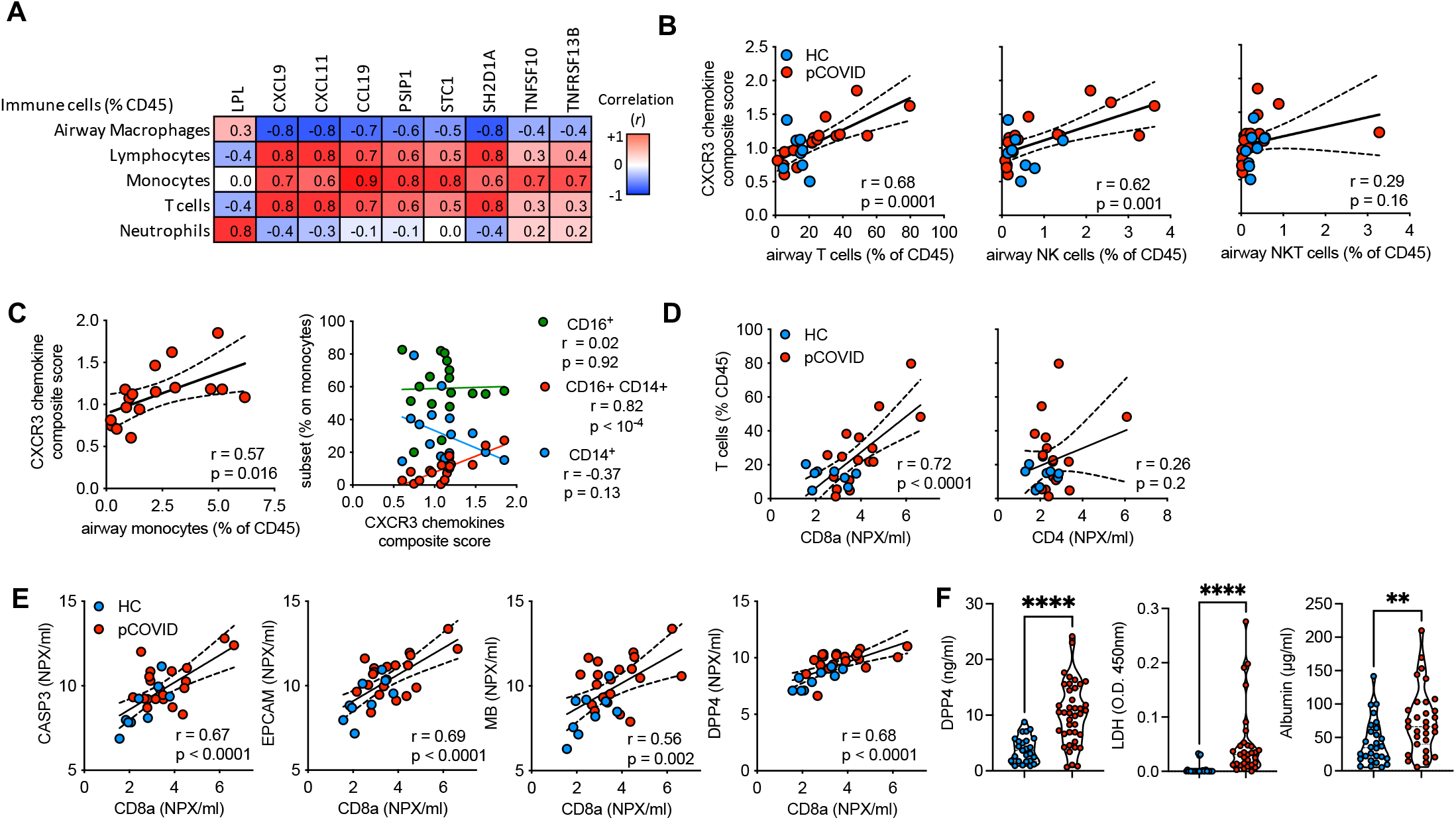
CXCR3 ligands and markers of epithelial damage correlate with CD8 T cells in the airways. BAL immune cells and protein concentrations were analysed post-COVID19 infection. **(A)** Linear regression analysis was conducted between n = 435 proteins measured in the BAL using the Olink platform and BAL immune cell frequencies identified by flow cytometry as shown in Figure 1. A heatmap showing proteins with the highest correlation versus 5 major immune cell frequencies is shown. **(B-C)** For each sample, protein levels for CXCL9, −10 and −11, were normalised to the median level in healthy controls. For each sample, the mean of the normalised values for the 3 proteins was calculated to provide a summary metric for CXCR3 chemokines. This was then plotted against versus (**B**) T, NK and NKT proportions in post-COVID19 patients and healthy controls and **(C)** monocyte frequencies and subsets in post-COVID19 patients only. **(D)** BAL T cell frequency versus CD4 and CD8a concentrations as measured by Olink. **(E)** CD8a concentration versus CASP3, EPCAM, MB and DPP4 in the airways. **(F)** DPP4, albumin and LDH concentrations in the BAL determined by ELISA. Data are presented as median ± IQR. **(A)** Pearsons correlation of n = 19 post-COVID19 patients, the r value is shown. **(B-E)** Each point represents an individual patient, linear regression line +/-95% confidence intervals are depicted, and r and p values from Pearsons correlation are stated. **(F)** represents n = 38 post-COVID19 and n = 20 healthy control individuals. Statistics were conducted using Mann-Whitney U test. *P < 0.05, **P < 0.01, ***P < 0.005, ****P < 0.001. pCOVID = post-COVID19.

Analysis of combined data from healthy controls and post-COVID19 patients revealed a composite score reflecting CXCL9, CXCL10 and CXCL11 levels, CXCL9, CXCL10 and CXCL11, although no significant correlation was seen with CD3^+^CD56^+^ NKT cells (**Figure 4B**). Within the post-COVID19 data set (as CD16 was not present in historic flow data used for healthy controls), total monocyte frequencies also correlated with average CXCR3 ligand expression (**Figure 4C**). Analysis of subpopulations revealed the proportion that were intermediate (CD14^+^CD16^+^) monocytes positively correlated with CXCR3 ligands, while CD14^+^ monocytes displayed a negative correlation, and CD16^+^ monocytes displayed no correlation (**Figure 4C**). T cell proportions in the airways correlated tightly with the concentration of CD8a protein, but not CD4, in the BAL (**Figure 4D**), suggesting the increased airway T cells are most likely the result of increased CD8^+^ T cell frequencies.

Given the robust correlation of the CXCR3 ligands, particularly with T cells, we next sought to determine the relationship between T cell frequencies and other members of the red module, specifically those indicative of ongoing epithelial damage in the post-COVID19 airways. CD8a correlated strongly with the concentrations of CASP3 and EPCAM, concomitant with two of the differentially expressed proteins: MB and DPP4 (**Figure 4E**). Collectively, these data suggest that proteins linked to the recruitment of T cells, especially cytotoxic T cells, are strongly associated with proteins that are both indicative of ongoing epithelial damage and upregulated in the airways post-COVID19.

To further evaluate this and confirm the presence of increased damage in the post-COVID19 airway, we measured DPP4, alongside 2 markers of damage not analyzed through Olink, albumin and lactate dehydrogenase (LDH) in an expanded cohort of healthy controls and post-COVID19 patients (**Figure 4F**). All 3 molecules were significantly upregulated in the airways of patients post-COVID19 compared to healthy controls, validating the observations made by Olink and confirming the presence of ongoing damage within the respiratory tract in patients previously hospitalized for COVID19.

### Distinct myeloid populations are present in the BAL compared to the peripheral blood but do not correlate with ongoing airway damage post-COVID19

Dysfunction of circulating classical monocytes is a hallmark of the initial, acute phase of COVID19 infection (Mann et al., 2020). Given the correlations observed between different airway monocyte populations and specific proteins in the BAL identified by Olink, we further investigated the nature of airway and circulating myeloid cells in a subgroup (n = 21) of post-COVID19 patients using high-parameter spectral deconvolution cytometry. Unbiased FlowSOM analysis of the airway and circulating myeloid cells revealed 10 distinct cell populations (**Figure 5A**), with marker analysis suggesting these populations could be assigned to known subsets of monocytes or dendritic cells (DCs). The circulation was dominated by CD14^+^ classical monocytes (metacluster 1), which constituted most myeloid cells identified, while the airways were more heterogenous, with the presence of intermediate monocytes, non-classical monocytes and DCs (**Figure 5B**). Conventional gating of these myeloid subsets (as shown in **Methods Figure 2**) confirmed these observations, with classical monocytes being most abundant in the circulation, while CD16^+^ monocytes were more frequent in the airways (**Figure 5C**). In comparison, DCs were identifiable in both sites as pDCs, cDC1s and cDC2s, although were infrequent (**Figure 5D**). Furthermore, airway monocyte activation status (as determined by CD86 and HLA-DR expression) was not dependent on acute disease severity (**Figure 5E**). Likewise, correlation of airway LDH and albumin concentrations revealed no strong associations between the relative abundance of different myeloid cells and ongoing damage in the respiratory tract after COVID19 (**Figure 5F**); nor was there any association between these cell types and DPP4 concentrations. Taken together this indicates that while myeloid cell frequencies in the airways are associated with a specific airway proteome landscape, this likely has limited association with ongoing tissue damage after SARS-CoV-2 infection.

**Figure 5.**
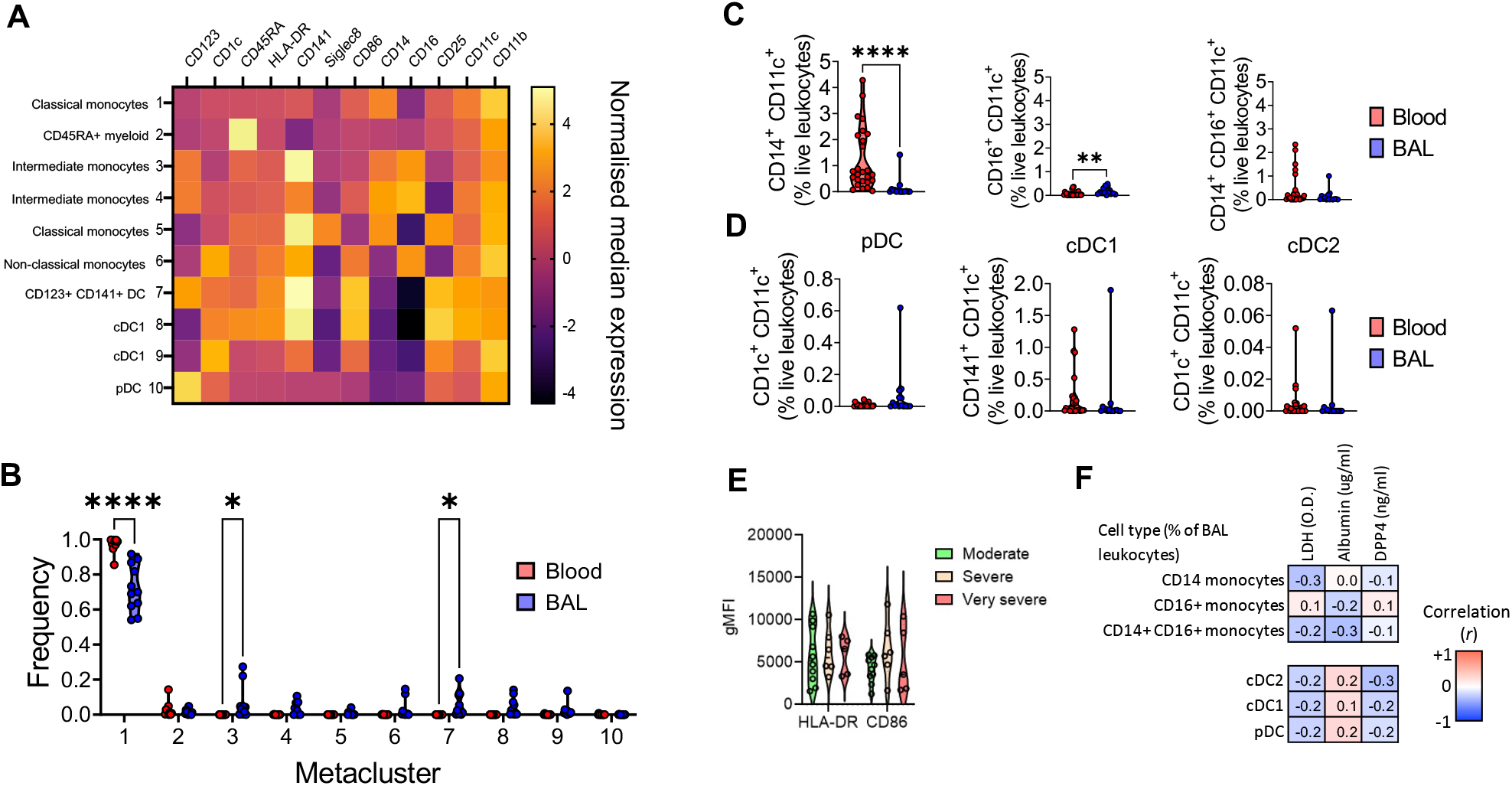
Myeloid cell frequencies are not linked to indications of ongoing damage in the post-COVID19 airway. BAL and blood immune cells from post-COVID19 patients were analyzed by spectral deconvolution cytometry. **(A)** Heatmap of normalised median expression of myeloid cell modulatory and subset markers by clusters of myeloid cells in the airways identified by FlowSOM analysis. **(B)** Violin plot showing frequencies of each cluster in BAL and blood. **(C)** Violin plots showing classical, non-classical and intermediate monocyte subsets as proportions of live leukocytes in BAL and blood identified by manual gating. **(D)** Violin plots showing pDC, cDC1 and cDC2 cell proportions in BAL and blood. **(E)** Geometric mean fluorescence intensity of activation markers expressed by CD11c^+^ monocytes. **(F)** Pearson’s correlation between BAL myeloid subsets (as a % of leukocytes) and airway LDH, albumin and DPP4. Data are presented as median ± IQR. Each point represents an individual patient. Statistical significance for **(B), (C)** and **(D)** was tested by Mann-Whitney U test. *P < 0.05, **P < 0.01, ***P < 0.005, ****P < 0.001.

### The post-COVID19 airway is enriched for activated, tissue resident T cells

The post-COVID19 airway showed significantly increased T cell numbers (**Figure 1C**), associated with a proteome linked to ongoing epithelial damage and repair (**Figure 2**). FlowSOM analysis of the airway and circulating lymphocytes indicated that 12 metaclusters with distinct protein expression profiles were present (**Figure 6A**). Several metaclusters were unique to either the blood or the airways, with specifically clusters 1 and 10 only present in the airways, and cluster 5 only present in the blood (**Figure 6B-C**). Marker expression analysis identified these metaclusters (as described in **Figure 6B**), with the differential clusters being identified as tissue-resident memory CD4 and CD8 T cells (metaclusters 1 and 10), and naïve CD4 T cells (metacluster 5). Manual gating of T cell populations showed that while the blood was enriched for naïve CD4 and CD8 T cells, the airways contained populations of antigen-experienced CD4 and CD8 T cells (**Figure 6D**). Additionally, the airways contained increased proportions of activated and tissue-resident memory (TRM) CD4 and CD8 T cells compared to blood (**Figure 6D**). Proportions of *γδ* T cells and NK cells in the airways and blood were comparable however there was a significant increase in NKT cell proportions compared to blood (**Figure 6E**). Comparison of the T cell dynamics in the post-COVID19 airways showed that airway CD4 T cells retain a highly tissue resident phenotype (CD69^+^CD45RA^-^), with few naïve (CD45RA^+^) CD4 T cells observed, and the frequency of these 2 cell populations remains relatively static, irrespective of the total number of CD4 T cells found in the airways (**Figure 6F**). In comparison, as CD8 T cell numbers increased in the airways, the proportion displaying a CD45RA^-^CD69^+^ phenotype significantly increased, while those with a CD45RA^+^ phenotype declined (**Figure 6F)**.

**Figure 6.**
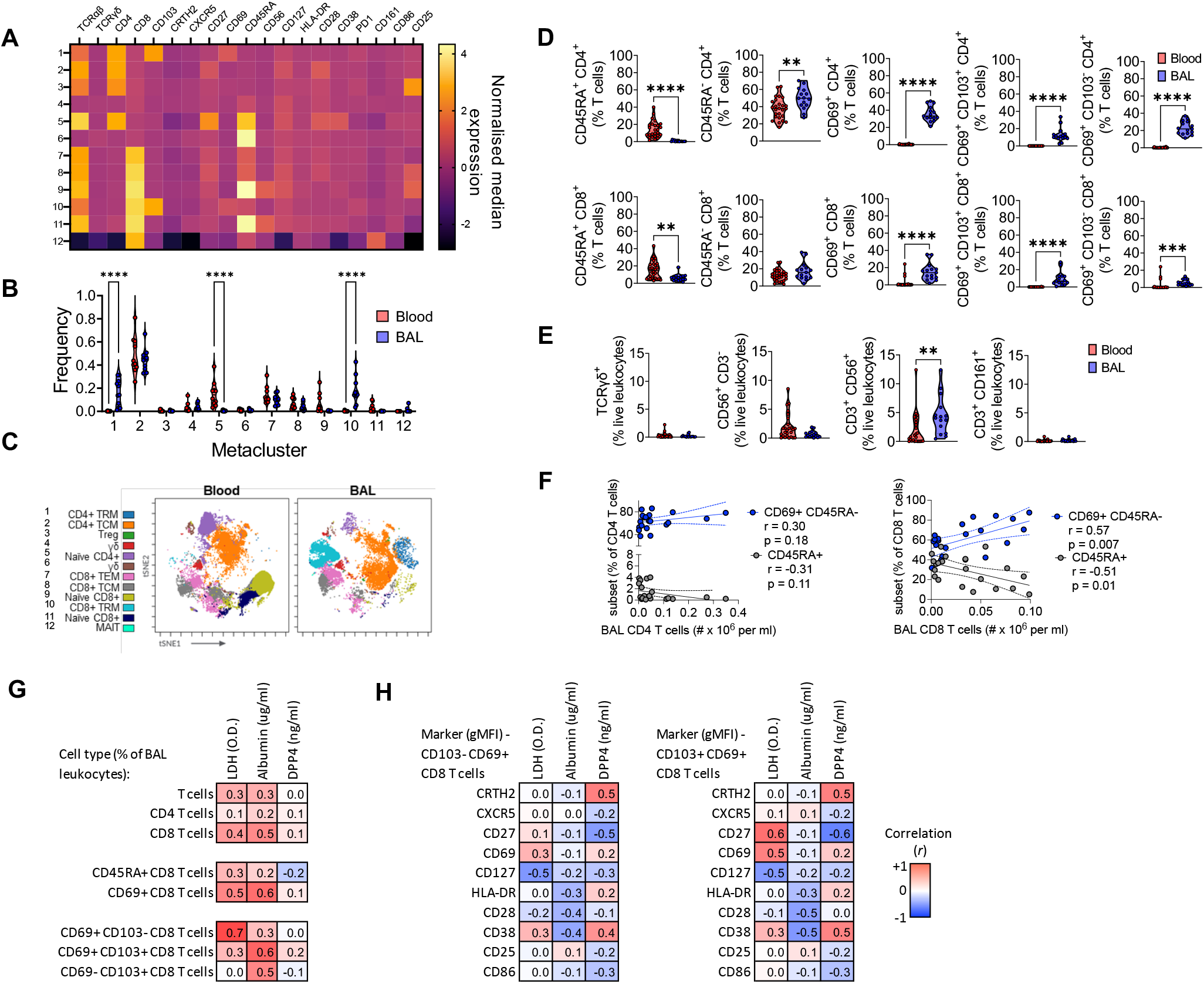
The airways of post-COVID19 patients are enriched for activated CD8 TRM cells. BAL and blood immune cells from post-COVID19 patients were analyzed by spectral deconvolution cytometry.**(A)** Heatmap of normalised median expression of T cell modulatory and subset markers by clusters of T cells in the airways identified by FlowSOM analysis. **(B)** Violin plot showing frequencies of each cluster in BAL and blood. **(C)** tSNE projection of clusters identified by FlowSOM analysis in BAL and blood. **(D)** Violin plots showing CD4+ and CD8+ T cell subsets as proportions of all T cells in BAL and blood identified by manual gating. **(E)** Violin plots showing gd, NK, NKT and MAIT cell proportions in BAL and blood. **(F)** Pearson correlations between total airway CD4^+^ and CD8^+^ T cell numbers and subsets. **(G)** Heatmap depicting Pearson correlation between different T cell populations (as a % of BAL leukocytes) and the concentration of DPP4, LDH and Albumin in the airways. **(F)** Pearson correlations between the gMFI of LDH and albumin. Data are presented as median ± IQR. Each point represents an individual patient. **(A, B, E** and **F)** represent n = 20 individuals. (**C**) represents an equal number of cells from n = 10 patients combined. (**G** & **H**) represents n = 18 individuals. Statistical significance for **(B), (D), (E)** and **(G)** was tested by Mann-Whitney U test. *P < 0.05, **P < 0.01, ***P < 0.005, ****P < 0.001.

In line with activated CD8 T cells, rather than activated CD4 T cells, being linked to ongoing epithelial damage in the post-COVID19 airway there was a correlation (r > 0.4) between CD8, but not CD4, T cell proportions in the airways and the concentrations of airway albumin and LDH (**Figure 6G**); this correlation was stronger when only CD69^+^ CD8 T cells were analyzed (r = 0.5, p < 0.05 for both Albumin and LDH). LDH most significantly correlated with the proportion of CD69^+^ CD103^-^ CD8 T cells in the airways (r = 0.7, p < 0.001), while albumin most significantly correlated with the proportion of CD69^+^ CD103^+^ CD8 T cells in the airways (r = 0.6, p < 0.001), highlighting distinct markers of damage were linked to different phenotypes of CD8 T cells in the airway. Of note however, the CD8 T cell frequency did not correlate with the concentration of DPP4 in the airways (**Figure 6G**). Deeper analysis revealed that albumin concentrations were associated with reduced expression of molecules associated with T cell activation on both CD103^+^ and CD103^-^ CD69^+^ CD8 T cells, most notable CD28 and CD38 (**Figure 6H**). LDH concentrations meanwhile were linked to downregulation of CD127 and, especially on CD103^+^ CD69^+^ CD8 T cells, but upregulation of CD69 and CD27 (CD69: r = 0.5, p < 0.05; CD27: r = 0.6, p < 0.05, **Figure 6H**). In contrast individuals with higher concentrations of DPP4 in the airways appeared to possess CD8 TRM with higher expression of CRTH2 and CD38 (**Figure 6H**). No relationship was seen between the expression of activation markers on the surface of CD8 TRM and the severity of disease in hospital (**Figure S5)**. Taken together, these results highlight the connection between T cells in the airways, specifically CD8 TRM, and ongoing epithelial damage after recovery from COVID19, and indicate differential CD8 T cell activation status may be associated with types of damage within the airways.

### The enhanced airway immune cell frequencies seen post-COVID19 decline over time

Acute COVID19 can result in persistent respiratory symptoms, and the data here highlight the presence of substantial immunological and proteome alterations in the airways of patients hospitalized with COVID19 up to 6 months after discharge. To determine if these changes to the airways reduced over time, we examined 3 patients with continued lung CT abnormalities greater than 1 year following discharge (**Table S3**). The total number of BAL cells recovered was greatly reduced in all 3 patients between the initial bronchoscopy and the 1 year follow up bronchoscopy, down to comparable levels in healthy control airways (**Figure 7A**). Similarly, numbers of T cells, B cells, NK cells and NKT cells were reduced to nearly or within the normal range seen in the airways of healthy individuals (**Figure 7B**). In the 2 individuals with elevated lymphocytes the ratio of CD4 to CD8 T cells increased (**Figure 7C**). In accordance with this the proportion of CD8, but not CD4, T cells trended to decrease, although the proportion of each that were of a TRM or activated (CD69^+^) phenotype, remained similar between the 2 time points (**Figure 7D**). Fitting with progressive recovery trajectory airway DPP4 concentrations declined in the 2 patients with elevated concentrations at the first bronchoscopy (**Figure 7E**), while CT abnormality was also reduced in all 3 patients between the first CT and the follow up (**Figure 7F**). Interestingly LDH was however increased between the first and second visit, while albumin concentrations were unchanged, but also within the range of healthy controls at both time points for all 3 patients analyzed (**Figure 7G**).

**Figure 7.**
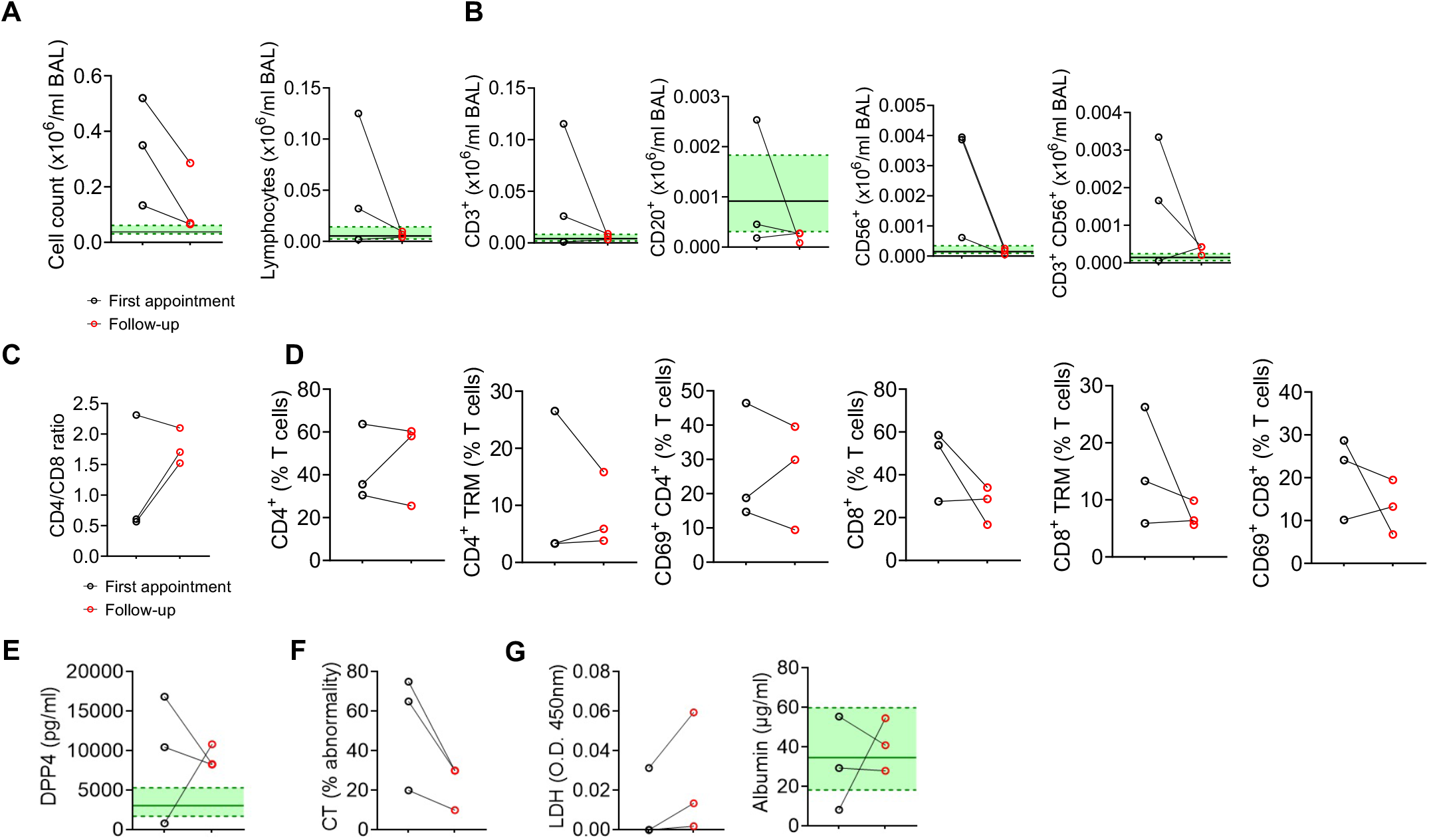
Reduced cellularity in the airways one year after initial bronchoscopy post-COVID19. **(A)** Total cell counts (left) and proportions of lymphocytes (right) in the BAL following first bronchoscopy and one year follow-up bronchoscopy. **(B)** Cell counts of lymphocyte populations in the BAL following first bronchoscopy and one year follow-up bronchoscopy. **(C)** Cell counts of myeloid populations in the BAL following first bronchoscopy and one year follow-up bronchoscopy. **(D)** Proportions of T cell subsets in the BAL following first bronchoscopy and one year follow-up bronchoscopy. **(E)** LDH (left) and albumin (right) measurements in BAL following first year bronchoscopy and one year follow-up bronchoscopy. **(F)** Percentage of abnormal CT following first year CT and one year follow-up CT. **(G)** BAL LDH and albumin quantification following first year bronchoscopy and one year follow-up bronchoscopy. Each point represents a single patient. Green shading indicates median+/-IQR for proportions of populations and mediator levels observed in healthy airways.

Collectively, our findings show ongoing changes to the immune and proteome landscape of the airways, characterized by increased T cells and markers of epithelial damage, several months after recovery from the acute phase of COVID19. These changes, and lung pathology, do however appear to resolve over the longer (> 1 year) term.

## Discussion

Recovery from acute SARS-CoV-2 infection is associated with a wide range of persistent symptoms including ongoing respiratory pathology. Here we examine the airway and circulating landscape of patients previously hospitalized with COVID19, finding they possess a distinct proteome and immunological profile in their airways, but not peripheral blood, 3 to 6 months post recovery from acute infection. While there is substantial heterogeneity between patients, common upregulated signals include proteins associated with ongoing cell death, epithelial damage and tissue repair; features that correlate with the presence of increased numbers of activated tissue resident CD8 T cells within the airways. Importantly, preliminary evidence suggests this altered airway landscape does improve over the long term, with reductions in airway immune cell numbers 1 year post discharge from hospital.

The acute response to SARS-CoV-2 infection is characterized by upregulation of a wide diversity of plasma proteins including members of the IFN pathway and their interferon stimulated genes (ISGs), chemokines, cytotoxic proteins, and markers of epithelial damage (Arunachalam et al., 2020; Filbin et al., 2021; Gisby et al., 2021). More severe disease is associated with increased concentrations of inflammatory proteins such as IL-6, TNF, GM-CSF, IL-1RN and IL-18 (Arunachalam et al., 2020; Filbin et al., 2021; Thwaites et al., 2021). A similar pattern of upregulated proteins, especially chemokines like CXCL10 and cytokines such as IL-6, is seen in the airways during acute COVID19 (Liao et al., 2020; Saris et al., 2021; Szabo et al., 2021). Here, we show that 3 to 6 months after SARS-CoV-2 infection, despite the presence of ongoing respiratory morbidity, the majority of the plasma proteins differentially expressed during acute disease appear to have returned to similar concentrations to those seen in healthy controls, and even data dimension reduction approaches such as WGCNA fail to highlight any significant associations between COVID19 infection and the plasma proteome months later.

In contrast, the post-COVID19 airways continue to display an abnormal proteome. This displayed both shared and distinct features to that seen in acute disease. Proteins linked to inflammation featured less prominently than in acute COVID19; instead, we observed airway upregulation of proteins involved in epithelial damage and repair (e.g. the EGFR ligand AREG and the epithelial marker KRT19). Matrix metalloproteinase-3 (MMP-3), which regulates the extracellular matrix (ECM), was also differentially upregulated in the post-COVID19 airway. MMP3 and AREG are both upregulated after influenza A virus (IAV) infection *in vivo* in mice, and *in vitro* in human fibroblasts and epithelial cells (Boyd et al., 2020); and these proteins are both linked to epithelial repair and fibrosis in the lungs e.g. (Morimoto et al., 2018; Yamashita et al., 2011).

The elevated concentrations of both lactate dehydrogenase and albumin in the airways provide further evidence of ongoing cell death and damage to the barrier integrity within the respiratory tract post-COVID19. This observation is reinforced by the upregulation of a module of proteins in the BAL of post-COVID19 patients whose individual members link epithelial damage (EPCAM, KRT19), cell death (CASP3) and epithelial repair (TGFA), but also suggest a connection between these processes and factors involved immune cell recruitment and survival (CXCL9, CXC10, CXCL11, IL-7). These markers of ongoing damage and repair tightly correlate with the frequency of T cells, especially CD8 TRM in the airways. In mouse models of severe acute respiratory virus infection CD8 T cells have long been known to act as a double-edged sword. Although the cytotoxic molecules and cytokines they release are essential for clearing virus, they can also cause tissue damage and immune pathology (reviewed in (Duan and Thomas, 2016; Schmidt and Varga, 2018)). While the presence of pre-existing virus specific CD8 TRM in the airways is thought to be highly protective against a re-encounter with the same virus in both mice and man (Jozwik et al., 2015; Wu et al., 2014) almost nothing is known regarding their role in long-term respiratory viral pathology, especially in humans. This is primarily due to the lack of relevant samples collected during the recovery period. However, the data shown here for SARS-CoV-2 may provide a potential insight, supporting the concept that sustained activation of CD8 TRM in the airways, long after recovery from acute disease, may contribute to ongoing immune pathology through sustained damage to the respiratory epithelium.

The mechanism behind the increased number of CD8 TRM in the airways is unclear, although another study has also reported detecting virus specific CD8 T cells in lung tissue up to a year post-infection (Grau-Exposito et al., 2021). Both virus specific CD4 and CD8 T cells rapidly expand in the circulation and airways of patients following SARS-CoV-2 infection, leading to the formation of TRM (Szabo et al., 2021). The majority of these activated effector T cells rapidly contract after resolution of acute disease. In mice Slutter *et al* have shown that lung CD8 TRM are more apoptotic than CD4 TRM after IAV infection, resulting in a more rapid decline in their number. They also observed that the lungs of a mouse that had previously experienced IAV infection more robustly maintained CD8 TRM compared to an uninfected lung, showing that severe infection promotes a pro-TRM niche (Slutter et al., 2017). This fits with our observation that while CD8 TRM numbers dynamically change dependent on the concentration of damage and proteins in the airways, and indeed longitudinally in the same individuals, CD4 TRM remain relatively static. A number of factors likely contribute to the heterogeneity of the CD8 TRM niche in the post-COVID19 airway. Firstly, while all our post-COVID19 samples were taken from patients who tested negative for SARS-CoV-2 by qPCR immediately prior to bronchoscopy, persistent antigen has been observed months after other respiratory infections such as IAV (Kim et al., 2010), and antigen depots in SARS-CoV-2 could drive ongoing cytotoxic activity and maintenance of CD8 TRM. Secondly, the persistence of lung resident TRM is also reliant on the availability of local T cell survival signals such as IL-7 (Szabo et al., 2019) and the CXCR3 ligands, which are known to be involved in airway recruitment and retention of CD8 memory T cells in murine IAV infection (Slutter et al., 2013). We found that both IL-7 and the CXCR3 ligands are part of the protein network that is maintained in the post-COVID19 airway. These proteins display a heterogeneous pattern of upregulation across post-COVID19 patient samples (**Figure 3B**) and correlate with airway T cells and CD8a concentrations. Lastly there is also some evidence of ongoing aberrant inflammation after acute infection and the development of autoantibodies in some patients recently recovered from COVID19 (Lucas et al., 2020; Wang et al., 2020). It is likely that these different mechanisms act in concert to shape the number and function of CD8 TRM, and other immune cells, in the post-COVID19 airway, and the scale and duration of ongoing epithelial damage and respiratory pathology observed.

One factor that does not appear to influence ongoing T cell recruitment and damage in the post-COVID19 airway is the activation status of the myeloid compartment, in particular monocytes. Functional impairment of monocytes and DCs in the peripheral blood of acutely infected patients (Arunachalam et al., 2020; Laing et al., 2020; Mann et al., 2020), and hyperactivation of airway monocyte populations, are canonical features of severe acute SARS-CoV-2 (Liao et al., 2020; Szabo et al., 2021). Moreover, early T cell phenotypes have been strongly linked to the status of these monocytes (Filbin et al., 2021; Laing et al., 2020; Szabo et al., 2021). In our post-COVID19 patients, however, the monocyte and myeloid pools in both the airways and peripheral blood appear to have returned to relatively normal numbers and their frequency does not correlate with indicators of ongoing epithelial damage. Likewise, their activation status, as determined by HLA-DR and CD86 expression, is not substantially different between individuals in line with other reports on circulating monocytes from earlier stages of convalescence (Scott et al., 2020).

The airway monocyte frequency is, however, one of the few features to correlate with a clinical biomarker of severity in acute disease, serum ferritin. High serum ferritin is a prognostic marker used at presentation with acute COVID19 and linked to more severe acute radiological findings (Carubbi et al., 2021; Ruan et al., 2020). Ferritin, and transferrin, the protein responsible for transporting ferritin, are core components of iron metabolism, and their dysregulation is associated with a range of lung pathologies including decreased lung function and lung fibrosis (Ali et al., 2020; Lee et al., 2020). In the respiratory tract the expression of CD71, the receptor that scavenges transferrin from the environment, on alveolar macrophages is altered in chronic respiratory disease (Striz et al., 1993), and the presence of CD71^-^ alveolar macrophages, possessing both an immature and pro-fibrotic transcriptional signature, has been linked to pulmonary fibrosis (Allden et al., 2019). In humans, monocytes are recruited to the airways to differentiate into new AMs (Byrne et al., 2020), and acute ferritin could be acting as a biomarker of severity of disruption to AMs. This link is however challenging to elucidate, and it is possible that ferritin is simply a proxy for the strength of the acute inflammatory response.

To our knowledge, this is the first study exploring the airway immune and proteomic profiles in COVID19 patients between 3 months and 1 year post discharge from hospital, and their links to ongoing respiratory pathology. Moreover, it is one of the first studies to explore the human airway immune-proteome landscape after a substantial period of time following any severe respiratory virus infection. Since all samples were obtained more than 3 months post hospital discharge, when one might expect normalization of the immune profile, the persistent immune abnormalities offer possible explanations into the longevity of persistent respiratory morbidity post SARS-CoV2 infection. In addition, these findings may be relevant to those suffering long-term sequalae during convalescence from other viral pneumonias where data are currently very limited.

We highlight a number of potential limitations of our study. Firstly, our data showing immunological and proteomic changes in the BAL of post-COVID19 are generated on patients undergoing clinically indicated bronchoscopy because of persistent respiratory abnormalities. Whether our findings extend to individuals with no radiological abnormalities or respiratory symptoms post-COVID19 remains unknown. This selection bias also affects longitudinal sampling greater than 12 months post-COVID19, since the majority of patients initially sampled between 3-6 months post-COVID19 had shown sufficient improvement in respiratory pathology such that a follow-up bronchoscopy was not indicated. The progressive resolution of radiological abnormalities in the majority of post-COVID19 patients has been observed by others (Han et al., 2021), and importantly within our study even the 3 patients with ongoing pathology have a significantly improved CT and reduced immune cell infiltration within their airways. This fits with the hypothesis that SARS-CoV2 infection can result in organizing pneumonia, with subsequent changes reflecting ongoing epithelial damage and healing parenchyma rather than established fibrosis (Kory and Kanne, 2020). This is both compatible with autopsy findings during the acute disease (Wichmann et al., 2020) and the steroid-responsive nature of the acute pathology (Horby et al., 2021).

Although we did not detect a plasma proteomic signature post-COVID19, this is likely due to our limited sample size not being powered to detect small differences in circulating proteins between post-COVID19 patients and healthy controls. Examination of the distributions of p-values suggests that such differences may exist (**Figure S3**) but will likely require future much larger cohort studies to reveal them. Regardless, the absence of any correlation between the differentially expressed proteins in the airways and their corresponding changes in the plasma points to the limited utility of peripheral blood as an indicator of the pathological processes ongoing in the lung.

Finally, as with most studies, we were limited to sampling the airways post-infection and so did not have paired pre-infection samples for intra-individual comparisons. Therefore, it is possible that some differences observed between healthy controls and post-COVID19 patients could reflect a pre-infection phenotype that predisposed them to developing prolonged sequelae. Indeed, one of the most differentially expressed proteins in the airways, DPP4, is the binding receptor for another coronavirus MERS (Raj et al., 2013), and postulated to be capable of mediating some SARS-CoV-2 binding (Li et al., 2020). Thus, it is conceivable that pre-existing upregulation of DPP4 increased susceptibility to post-COVID19 syndrome via increased viral entry (i.e. reverse causation), rather than DPP4 upregulation occurring in response to COVID19. However, the longitudinal reduction of DPP4 argues against this hypothesis. More generally, the majority of proteins and markers upregulated are associated with ongoing lung pathology in other contexts (e.g. LDH), and are absent or only present at very low concentrations in the healthy airway, suggesting that their upregulation is more likely to be a consequence of COVID19 than a pre-disposing risk factor.

In summary, our study offers unique and novel insights into ongoing immunopathology post-COVID19. In contrast to the inflammatory pathways observed during acute disease, we found proteins associated with ongoing epithelial damage, cell death, and repair were upregulated in conjunction with ongoing CD8 T cell activation. These data indicate that significant immune pathways operate within the tissue, presumably to facilitate repair and resolution, even in the absence of peripheral inflammation. In the future it will be important to determine how such a substantial shift in the immune landscape of the airways might affect the response to a subsequent respiratory infection such as seasonal influenza. The progressive improvement in respiratory pathology, even in this cohort with substantive radiological abnormalities should be seen as a positive indicator for the long-term prognosis for those with persistent morbidity. Moreover, the involvement of the immune response suggests this recovery could be accelerated using immuno-modulatory treatments, especially those designed against TH1 immune responses.

## Supporting information

supplemental data sheet

## Data Availability

All data will be made available for this study upon acceptance after peer review.

## Acknowledgements

We thank Jack Gisby for statistical and analytical advice and Simone Walker for technical assistance. We also thank the staff of Sir Alexander Fleming Building Flow Cytometry Facility at Imperial College London for assistance with flow acquisition and analysis. The schematic in Figure 1A was made using Biorender. We also thank Dr Ryan Thwaites and Professor Peter Openshaw, Imperial College London, for critically reading this manuscript before submission.

## Author contributions

**BV** and **PLS** designed the PHENOTYPE study and were involved in recruitment of the post-COVID19 cohort. **PLS, CML, JEP** and **JAH** designed and conceived the current study. **JAH** and **CML** co-ordinated the study. **BV, JT, CO, JG** and **PLS** collected clinical samples and patient information for the post-COVID19 cohort. **RJH** and **PLM** designed and were responsible for recruitment and sampling of the heathy control cohort. **BV, KB** and **PPO** processed clinical samples for the post-COVID19 cohort. **PG** processed clinical samples for the heathy control cohort. **BV, KB, PPO** and **PG** conducted flow cytometry and mediator measurements. **KS** measured LDH and albumin. **KB** and **PPO** analysed the flow cytometry analysis. **AP** analyzed the Olink proteomics data set. **PS** supervised the clinical aspects of the project, **JEP** supervised the bioinformatic analysis. **RJS, PLM, CML** and **JAH** supervised the laboratory preparation of samples and flow cytometry and mediator measurement analysis. **BV, KB, PPO, AP, JEP, PLS, CML** and **JAH** were involved in data interpretation. **JAH** wrote the initial draft of the manuscript and **BV, KB, PPO, AP, JEP, PLS** and **CML** carried out major writing of the paper. All the authors reviewed and approved the submitted manuscript. For information on proteomics contact James E Peters; For clinical information contact Pallav L Shah; For epithelial cell biology and repair contact Clare M Lloyd; For immune cell phenotyping contact James A Harker.

## Methods

### Post-COVID-19 patient recruitment

Post-COVID19 bronchoalveolar lavage fluid (BAL) was obtained from patients recruited to the PHENOTYPE study (NCT 04459351), an observational, longitudinal study recruiting patients at Chelsea and Westminster Hospital, London. 38 samples were collected from patients requiring sampling for clinical purposes. Ethical approval for the study was given by Yorkshire & The Humber - Sheffield Research Ethics Committee (IRAS 284497).

Patients who met the inclusion and exclusion criteria were recruited to the PHENOTYPE study:

Inclusion criteria for the study were:

- Aged 18 years or older
- Previous confirmed COVID-19 infection (positive PCR or antibody)
- Attending a respiratory follow-up outpatient appointment for follow-up of persistent respiratory symptoms following visit post hospital attendance with COVID-19. infection or referred by the community for covid-related symptoms.

Patients were seen at approximately 4-6 weeks (Visit 1) and 3 months (Visit 2) following discharge from hospital or referral (if referred from the community). Patients underwent clinical assessment at both visits, including collection of demographic data, clinical history and clinical examination and assessment of vital parameters (heart rate, peripheral oxygen saturations, blood pressure reading and temperature). They also underwent clinical blood tests (including full blood count, renal function, liver function, C-reactive protein (CRP), ferritin, fibrinogen, D-dimer and pro-calcitonin). Patients had a computed tomography (CT) scan of the lungs approximately 3 months post discharge from hospital. In patients with abnormal CT findings, or persistent respiratory symptoms, a bronchoscopy and lavage was performed as part of clinical work-up. Patients underwent formal lung function tests (including spirometry, lung volumes and gas transfer) near the time of the bronchoscopy (usually during the days immediately preceding the procedure). Further follow-up was determined on the basis of clinical need, with a maximum follow up period of up to 2 years post hospital discharge or referral.

### Post-COVID19 bronchoalveolar lavage (BAL) sampling

Bronchoscopy was performed under conscious or deep sedation. 150 ml of normal saline were instilled into the most affected segment (as determined by CT imaging), in 50 ml aliquots. 10 ml of fluid return was used for the scientific analysis described in this paper.

### Healthy volunteer recruitment and sampling

Control, uninfected bronchoalveolar lavage was obtained from healthy donors (collected between April 2016 and December 2019). Ethical approval for the study was granted by the Research Ethics Committee (15/SC/0101) and all patients provided informed written consent as described previously (Allden et al., 2019; Byrne et al., 2020; Invernizzi et al., 2021). Briefly, 240 ml aliquots of warmed sterile saline were instilled in the right middle lung and aspirated by syringe. Lavage aliquots were pooled for each subject. All subjects provided written, informed consent to participate in the study. Healthy volunteers had no self-reported history of lung disease, an absence of infection within the last 6 months and normal spirometry.

### BAL processing

BAL samples were processed and stained on the day of sample collection. BAL was strained through a 70μm filter and subsequently centrifuged (1800 rpm, 2 min, 4°C). Supernatant was snap-frozen and stored at −80°C. Pellets were incubated in red blood cell lysis buffer (155mM NH_4_Cl, 10mM KHCO_3_, 0.1mM ethylenediaminetetraacetic acid, pH 7.4) for 10 minutes before washing and resuspension in complete media (RPMI 1640 with 10% fetal calf serum, 2mM L-glutamine, 100U/ml penicillin-streptomycin).

### Blood processing

Peripheral blood was collected in EDTA coated vacutainers on the same day as bronchoscopy. 1ml blood was centrifuged at 100g for 10 minutes (4°C), followed by centrifugation at 20,000g for 20 minutes (4°C) to separate plasma, which was subsequently stored at −80°C. 2ml blood from post COVID-19 patients was incubated with red blood cell lysis buffer (155mM NH_4_Cl, 10mM KHCO_3_, 0.1mM ethylenediaminetetraacetic acid, pH 7.4) for 10 minutes before washing and resuspension in complete media (RPMI 1640 with 10% fetal calf serum, 2mM L-glutamine, 100U/ml penicillin-streptomycin). 2.5 ml blood from healthy controls was used to isolate peripheral blood mononuclear cells (PBMC) by Percoll density centrifugation, as per manufacturer’s instructions.

### Flow cytometry staining

For traditional flow cytometry, 2 - 5 ⨯10^5^ cells were plated, while for high parameter analysis using the Cytek Aurora 1 ⨯ 10^6^ cells from each site were used. Cells were washed with PBS and incubated with either near-infrared (traditional flow cytometry) or blue (Cytek Aurora) fixable live/dead stain (Life Technologies), as per the manufacturer’s instructions. Before incubation with human fc block (BD Pharmingen) cells were washed with FACS buffer (1% FCS, 2.5% HEPES, 1mM EDTA) and surface staining was performed at 4°C for 30 minutes using antibody panels as described in the **Key Resources Table**. Surface staining was followed by washing with FACS buffer and fixation with 1% paraformaldehyde for 10 minutes. Labelled cells were acquired on a 4-laser BD Fortessa (traditional flow cytometry; BD Bioscience) or 5-laser Cytek Aurora flow cytometer (Cytek Bio).

### Flow cytometry analysis

Conventional flow cytometry data was analysed using FlowJo v 10.6 (Tree Star). Data was pre-gated to exclude doublets and dead cells. In BAL samples CD45^+^ cells were selected, and immune cell populations were identified using the gating strategy shown in **Methods Figure 1**. Percentages of the CD45^+^ gate were calculated. In blood samples, leukocytes were selected based on FSC and SSC and immune cell populations were identified using the gating strategy shown in **Methods Figure 1**. Percentages of total leukocytes were calculated. High-parameter spectral deconvolution flow cytometry data from the Cytek Aurora was analysed using Cytobank (Beckman). tSNE analysis was performed on 300,000 events from 11 files. Iteration number was set to 1500 with a perplexity of 30 and theta of 0.5. FlowSOM analysis was performed subsequently using hierarchical consensus clustering with 12 metaclusters, 100 clusters and 10 iterations. Manual gating of high parameter cytometry data was carried out as shown in **Methods Figure 2**. Heatmaps were generated from median fluorescence values in Prism 9.0 (GraphPad).

### Quantification and statistical analysis for flow cytometry

Differences in means between two sample groups were compared using two-tailed Mann-Whitney U tests. Multiple group comparisons were done using Kruskal Wallis ANOVA followed by Dunn’s post-test. Spearman-Rank correlations were used to correlate clinical blood parameters with immune cell populations. Analysis was performed in GraphPad Prism. For all figures, * denotes p value < 0.05, ** denotes p value < 0.01 and *** denotes p value < 0.001.

### Proteomic assays

Plasma and BAL proteomic measurement was performed using the Olink proximity extension immunoassay platform. Five 92-protein multiplex Olink panels were used (‘Inflammation’, ‘Immune Response’, ‘Cardiometabolic’, Cardiovascular 2’, ‘Cardiovascular 3’), providing measurements of 460 protein targets per sample. Cryopreserved BAL and plasma samples were thawed on ice and mixed well by pipetting before plating 88 samples per plate ensuring case/control balance and random well ordering to prevent confounding of technical and biological effects. For BAL samples, a pilot study was performed using three control samples and three post-COVID19 samples (severe group) to determine optimal dilution parameters. Ultimately BAL was used neat. Since a small number of proteins were assayed on more than one panel, we measured a total of 435 unique proteins. We removed duplicate assays at random prior to subsequent analyses.

### Proteomics analyses

Proteomic data was normalised using standard Olink workflows to produce relative protein abundance on a log2 scale (‘NPX’). BAL and plasma proteomic data were normalised separately. Quality assessment was performed by (1) examination of Olink internal controls and (2) inspection of boxplots, relative log expression plots, and PCA.

PCA was performed using singular value decomposition. Following these steps, 2 clear outlying samples were removed from the BAL dataset.

To identify proteins that were differentially abundant between case and controls, for each protein we performed linear regression (lm function in R) with case/control status as the independent variable and protein level (NPX) as the dependent variable. P-values were adjusted for multiple testing using the Benjamini-Hochberg procedure (p.adjust function in R). A 5% false discovery rate was used to define statistical significance.

We used the WGCNA R package (Langfelder and Horvath, 2008; Zhang and Horvath, 2005) to create a weighted protein correlation network. Prior to WGCNA analysis, protein data were scaled and centred, and missing data were imputed using the R caret package. We used the WCGNA adjacency function to produce a weighed network adjacency matrix, using parameters “type=signed” and “power=13”. This soft-thresholding power was selected as the lowest power to achieve approximate scale-free topology. We next defined a topological overlap matrix of dissimilarity using the TOMdist function. Clusters (‘modules’) of interconnected proteins were identified using hierarchical clustering and the cutreeDynamic function with parameters: method=“hybrid”, deepSplit=2, minClusterSize=15. We then tested association of these modules with case/control status. Multiple testing correction was performed to account for the number of modules. We report both Benjamini-Hochberg and Bonferroni adjusted p-values to provide two levels of stringency.

To assess the distribution of p-values from the differential protein abundance analyses, we plotted histograms and constructed QQ plots. QQ plots were made by comparing the expected distribution of −log10 P values under the null hypothesis of no proteomic differences between post-COVID19 patients and controls to the observed p-values for the 435 proteins.

We performed pathway enrichment analysis for the 435 proteins measured. This was performed using terms from KEGG database (**Supplementary File 1B**) and the Reactome database (**Supplementary File 1C**).

Protein modules were visualised using STRING (https://string-db.org/), with known or suspected interconnections between module members displayed as edges in a network diagram. An edge represents a protein-to-protein relationship defined as shared contributions to a particular function, and not necessarily implying physical binding. In **Figure 3C**, edge colour indicates the type of evidence for the relationship: turquoise represents known interactions from curated databases; magenta represents experimentally determined interactions; green represents predicted Interactions from gene neighbourhood analysis; red represent predicted interactions from gene fusions, blue represent predicted Interactions from gene co-occurrence; light green represents interaction from text-mining; black represents interaction from co-expression data, and violet represents information from protein homology.

### CXCR3 chemokine composite score

To create a composite score that reflected the CXCR3 chemokines (CXCL9, CXCL10 and CXCL11), we used the following approach. For each sample, protein levels for CXCL9, −10 and −11, were normalised to the median level in healthy controls (to avoid unduly weighting the score towards chemokines with higher NPX values). For each sample, the mean of the normalised values for the 3 proteins was then calculated to provide a summary metric for CXCR3 chemokines.

### Quantification of mediators in BAL

DPP4 (R&D systems, DY1180) and albumin (Bethyl Laboratories, E80-129) concentrations in the BAL were quantified by ELISA according to manufacturer’s instructions. LDH concentrations were quantified using an *in vitro* toxicology assay (Sigma, TOX7). Briefly, 25μl of BAL sample were incubated with 50μl of LDH assay reaction mixture. After 30 minutes, the reaction was stopped with 7.5μl 1N HCL and absorbance was measured at 490nm with background correction at 690nm. All absorbances were measured using a SpectraMax i3x (Molecular Devices).

### Key Resources Table

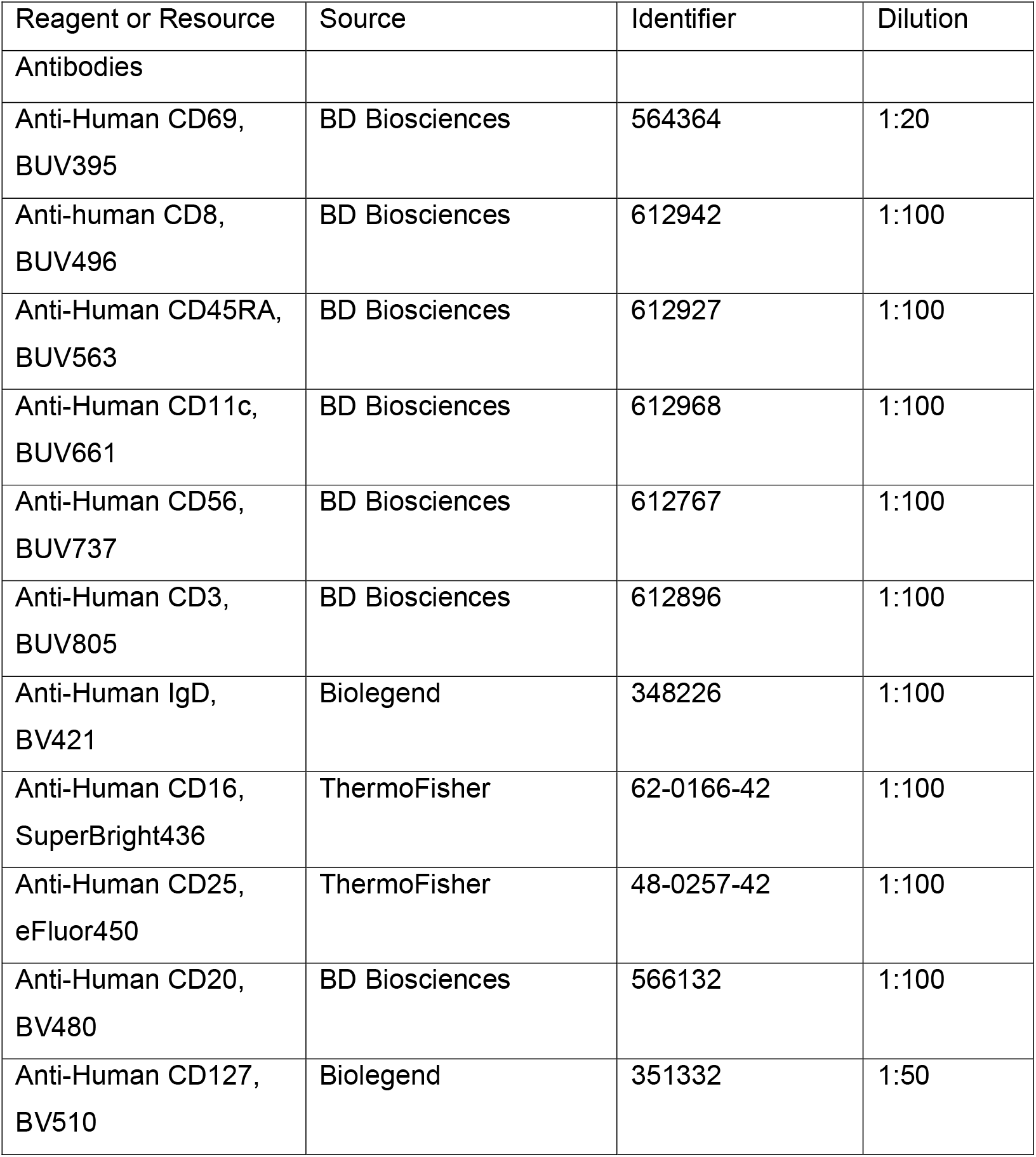
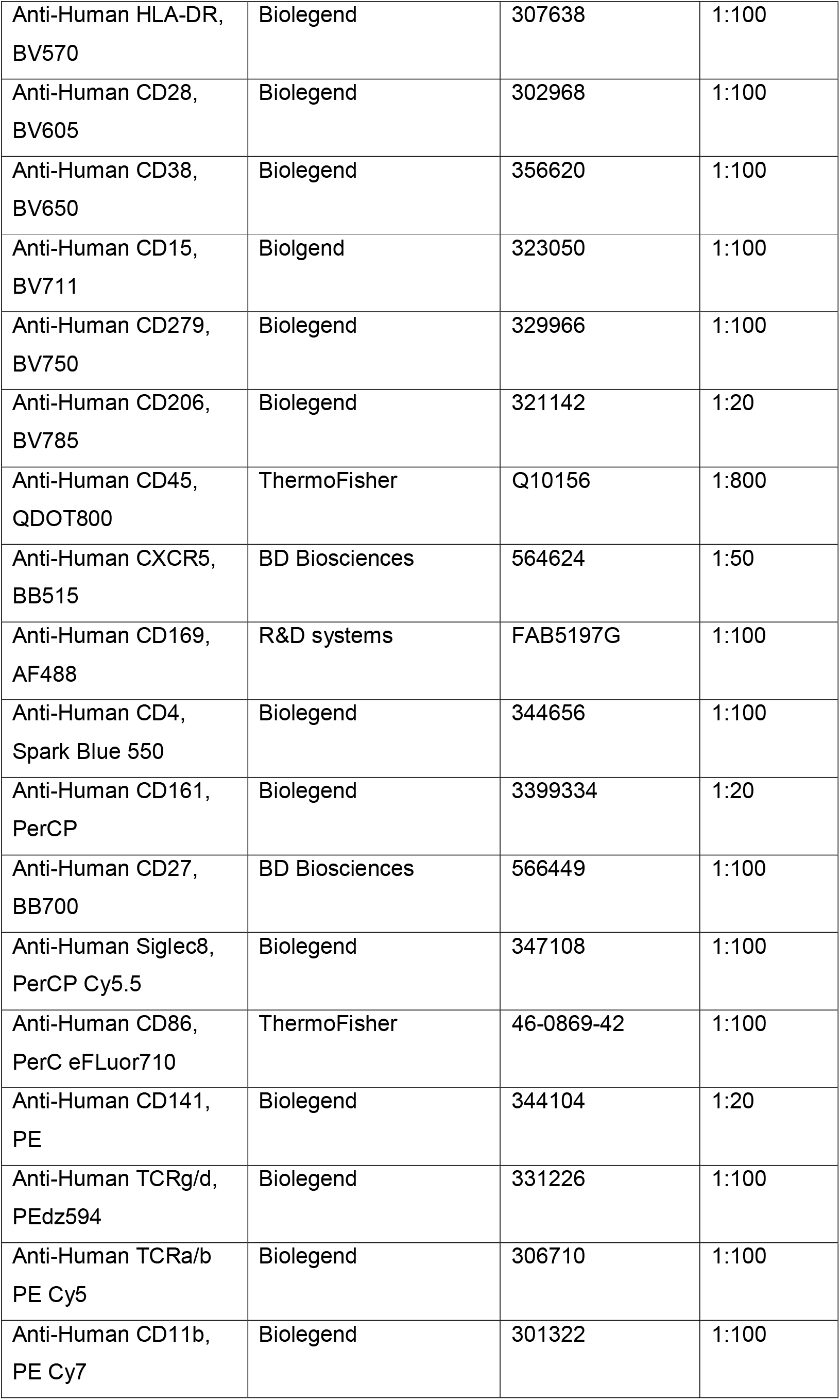
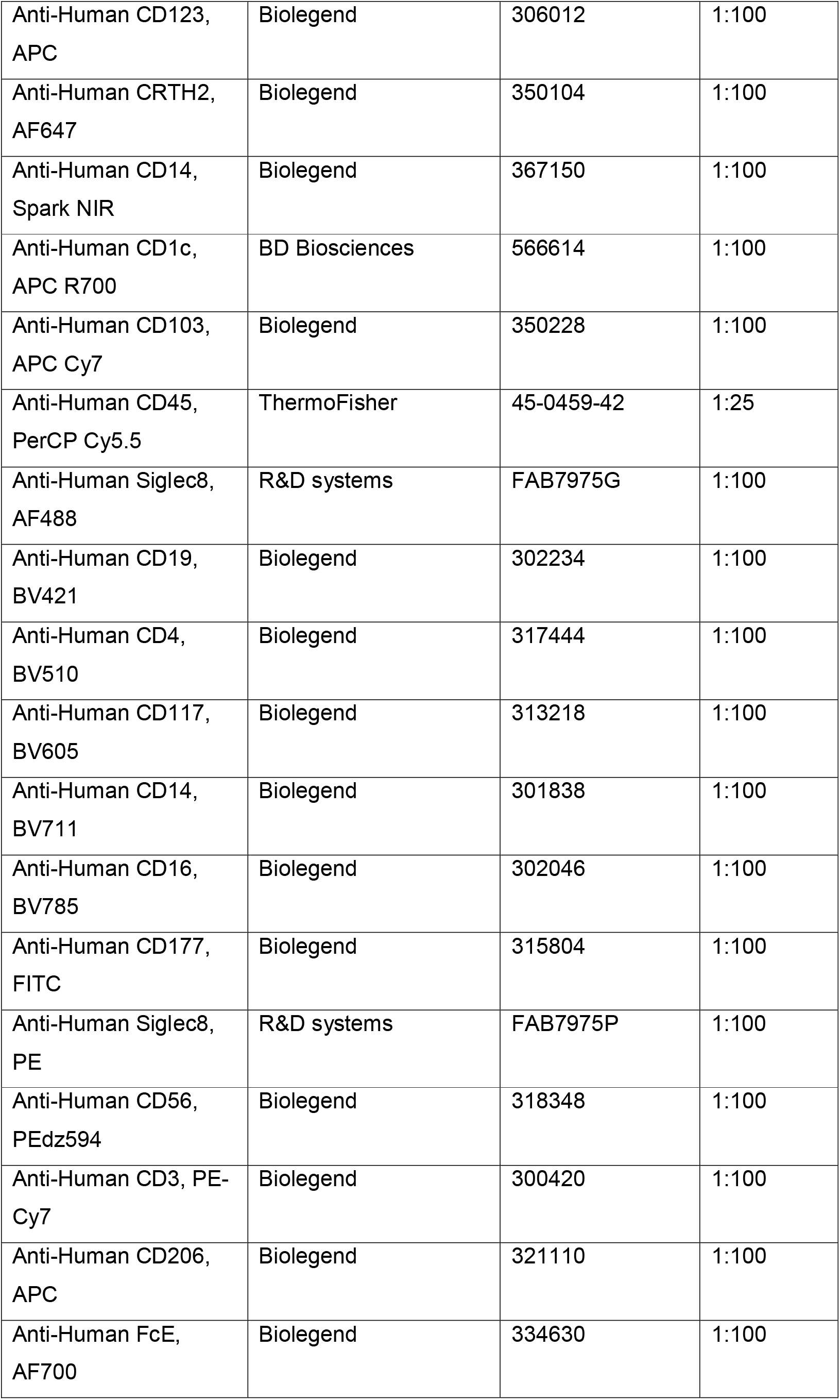
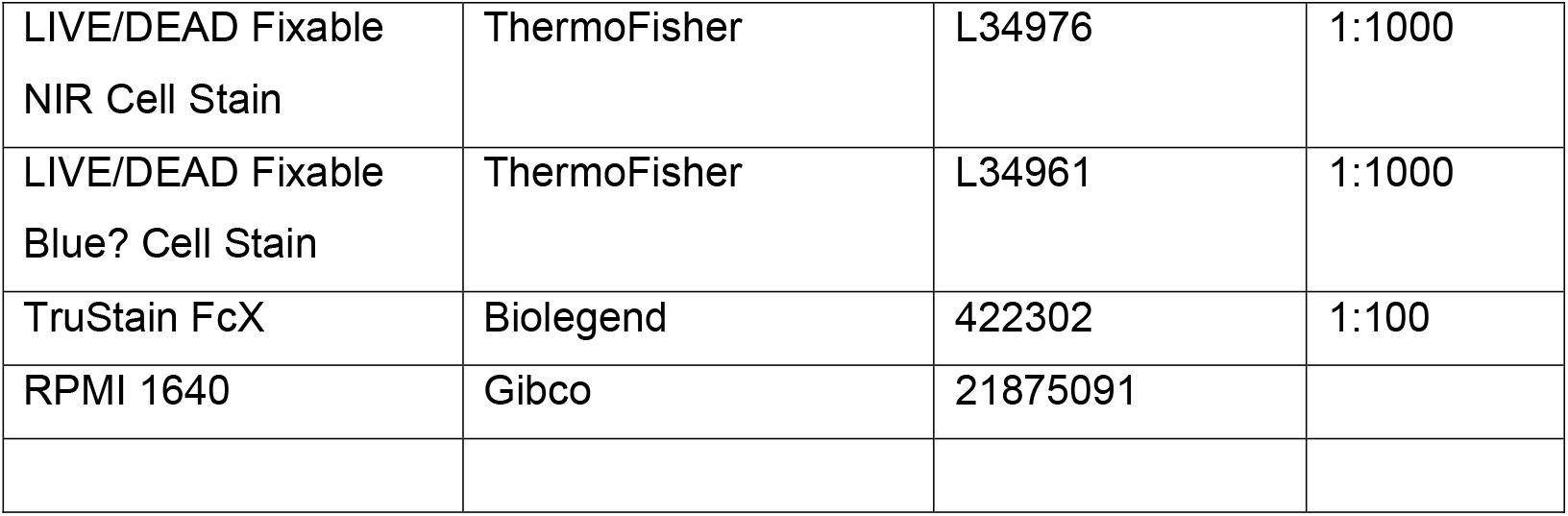

