## Supplemental tables and figures for "Immuno-proteomic profiling reveals abundant airway CD8 T cells and ongoing epithelial injury in prolonged post-COVID19 respiratory disease"

**Table S1: Demographics of the post-COVID19 and healthy control cohort**

Abbreviations: Gender, M (male), BAME (black, Asian and minority ethnic), COPD (chronic obstructive pulmonary disease), CPAP (continuous positive airway pressure), NIV (Non-invasive ventilation), IMV (invasive mechanical ventilation), CT (computed tomography), FEV1 (forced expiratory volume in 1 second), FVC (forced vital capacity), TLC (total lung capacity), TLCO (transfer factor of the lung for carbon monoxide), KCO (carbon monoxide transfer coefficient. Data presented as n (%n), median (IQR)

| | Post-COVID19 (n = 38) | Healthy controls (n = 29) |
|---|---|---|
| Age (years) | 56.5 (51.3-66.0) | 49.0 (39.5 - 54.5) |
| Gender, M (%n) | 30 (78.9) | 17 (58.6) |
| BAME (%n) | 22 (57.8) | N/A |
| Comorbidities (%n) | | |
| Asthma | 5 (13.2) | N/A |
| COPD | 1 (2.63) | N/A |
| Previous pneumothorax | 1 (2.63) | N/A |
| Hypertension | 9 (23.7) | N/A |
| Ischaemic heart disease | 3 (7.89) | N/A |
| Type 2 diabetes mellitus | 7 (18.4) | N/A |
| Smoking history | | |
| Current smoker | 1 (2.63) | 2 (6.8) |
| Ex-smoker (>=10 pack year) | 5 (13.2) | 4 (13.7) |
| Non-smoker or ex-smoker <10 pack year) | 32 (84.2) | 17 (58.6) |
| Severity of acute illness | | |
| Hospitalized (n/%) | 36 (94.7) | N/A |
| - Oxygen therapy only (moderate) | 15 (39.5) | N/A |
| - CPAP/ NIV (severe) | 11 (28.9) | N/A |
| - IMV (very severe) | 10 (26.3) | N/A |
| Not hospitalized (moderate) | 2 (5.26) | N/A |
| Length of hospital stay (days) [n=35] | 10.0 (5.0-27.0) | N/A |
| Admission bloods [n=33] | | |
| White cell count ($x10^9$/L) [n=33] | 6.80 (5.70-8.85) | N/A |
| Haemaglobin (g/L) [n=33] | 143 (132-154) | N/A |
| Platelet ($x10^9$/L) [n=33] | 219 (195 – 271) | N/A |
| Neutrophil ($x10^9$/L) [n=33] | 5.90 (3.75-7.30) | N/A |
| Lymphocyte ($x10^9$/L [n=33] | 1.0 (0.70 – 1.45) | N/A |
| D dimer (microgram/L) [n=25] | 965 (800-1622) | N/A |
| Fibrinogen (g/L) [n=31] | 7.23 (6.21-8.09) | N/A |
| Ferritin (micrograms/L) [n=26] | 762 (606-2329) | N/A |
| Sodium (mmol/L) [n=33] | 134 (133-137) | N/A |
| Potassium (mmol/L) (n=31) | 4.30 (4.1– 4.70) | N/A |
| Urea (mmol/L) [n=33] | 5.30 (3.40-7.10) | N/A |
| Creatinine (micromol/L) [n=33] | 86.0 (75.0-103) | N/A |
| Egfr (ml/min/1.73m2) [n=33] | 79.0 (58.0-90.0) | N/A |
| Albumin (g/dL) [n=33] | 33.0 (30.5-34.0) | N/A |
| C-reactive protein (mg/L) [n=33] | 124 (84.1 – 223) | N/A |
| CT imaging (n=38) | | |
| Days from admission to follow up CT [n=35] | 117 (93.0-141) | N/A |
| Days from discharge to follow up CT [n=36] | 97 (84.3-127) | N/A |
| Abnormal CT | | N/A |
| Overall CT abnormality | | N/A |
| Ground glass abnormality | | N/A |
| Reticulation | | N/A |
| Consolidation | | N/A |
| Bands | | N/A |
| Bronchoalveolar lavage fluid sampling (n=38) | | |
| Days between admission and sampling [n=35] | 153 (107-192) | N/A |
| Days between discharge and sampling [n=36] | 135 (98.5-176) | N/A |
| Lung function tests [ n=36] | | |
| Days between admission and lung function tests [n=33] | 147 (106-185) | N/A |
| Days between discharge and lung function tests [n=34] | 118 (92.0-171) | N/A |
| FEV1 (L) | 2.97 (2.36-3.60) | 3.26 (2.8 - 3.6) |
| % predicted FEV1 | 95.5 (86.5-103) | |
| FVC (L) | 3.73 (2.83 – 4.16) | 3.96 (3.5 – 4.9) |
| % predicted FVC | 91.5 (83.5 – 96.8) | |
| TLC (L) | 5.73 (5.09 – 6.33) | N/A |
| % predicted TLC | 89.0 (83.3 – 99.8) | N/A |
| TLCO ml/min/mmHg | 6.39 (5.32 – 8.57) | N/A |
| % predicted TLCO | 78.5 (67.3 – 96.8) | N/A |
| KCO ml/min/mmHg | 1.37 (1.20-1.61) | N/A |
| % predicted KCO | 99.0 (88.3-115) | N/A |

| BAL | % CD45 | | | | | | total cell numbers | | | | | |
|---|---|---|---|---|---|---|---|---|---|---|---|---|
| highest value during hospitalisation | WCC | D dimer | Fibrinogen | Ferritin | CRP | Lymphocytes | WCC | D dimer | Fibrinogen | Ferritin | CRP | Lymphocytes |
| AM | -0.13 | -0.08 | 0.08 | -0.44 | 0.16 | 0.06 | 0.08 | 0.22 | 0.36 | -0.19 | 0.02 | 0.2071 |
| CD14+ Monocytes | 0.04 | -0.06 | -0.25 | 0.62 | -0.12 | -0.12 | 0.07 | 0.10 | -0.03 | 0.53 | -0.21 | -0.0333 |
| CD14+, CD16+ Monocytes | 0.05 | 0.16 | -0.24 | 0.58 | -0.29 | -0.22 | 0.01 | 0.20 | -0.17 | 0.50 | -0.41 | -0.1852 |
| CD16+ Monocytes | -0.16 | 0.19 | -0.33 | 0.25 | -0.25 | -0.02 | -0.03 | 0.18 | -0.17 | 0.30 | -0.38 | 0.07909 |
| total Monocytes | -0.14 | 0.09 | -0.37 | 0.48 | -0.26 | -0.10 | -0.10 | 0.05 | -0.17 | 0.45 | -0.44 | 0.05411 |
| T cells | -0.04 | 0.17 | 0.01 | 0.25 | -0.18 | -0.02 | 0.06 | 0.29 | 0.14 | 0.25 | -0.15 | 0.1051 |
| NK T cells | 0.34 | 0.02 | 0.14 | 0.38 | 0.01 | -0.34 | 0.25 | 0.06 | 0.12 | 0.39 | -0.19 | -0.1739 |
| B cells | 0.07 | 0.49 | 0.30 | 0.27 | 0.30 | 0.06 | 0.06 | 0.50 | 0.35 | 0.26 | 0.30 | 0.1249 |
| NK cells | -0.01 | 0.13 | 0.13 | 0.13 | 0.11 | 0.15 | 0.05 | 0.29 | 0.24 | 0.12 | 0.02 | 0.2258 |
| Neutrophils | 0.24 | -0.29 | -0.32 | 0.14 | -0.23 | -0.31 | 0.17 | -0.28 | -0.22 | 0.26 | -0.36 | -0.1499 |
| Eosinophils | -0.07 | -0.05 | -0.12 | 0.40 | -0.11 | 0.42 | -0.18 | 0.00 | -0.10 | 0.30 | -0.27 | 0.4912 |
| Lymphocytes | 0.00 | 0.21 | 0.03 | 0.30 | -0.12 | -0.04 | 0.07 | 0.32 | 0.14 | 0.36 | -0.13 | 0.08325 |

**Table S2: Spearman Rank correlation between blood parameters during hospitalisation and immune cell populations in BAL at > 80 days post discharge**

R values of Spearman Rank correlation between blood parameters and immune cell populations in BAL from pCOVID patients. WCC, D-dimer, fibrinogen, ferritin and CRP values are the highest recorded value during hospitalisation. N = 19; Values in green are R > 0.4, values in blue are R < -0.4.

**Table S3.** Demographics, blood tests, CT abnormalities, BALF findings and lung function tests at initial and follow bronchoscopy.
Abbreviations: Gender, M (male), BAME (black, Asian and minority ethnic), COPD (chronic obstructive pulmonary disease), CPAP (continuous positive airway pressure), NIV (Non-invasive ventilation), IMV (invasive mechanical ventilation), CT (computed tomography), FEV1 (forced expiratory volume in 1 second), FVC (forced vital capacity), TLC (total lung capacity), TLCO (transfer factor of the lung for carbon monoxide), KCO (carbon monoxide transfer coefficient
Data presented as n (%n), median (IQR)

| | pCOVID (n = 3) |
|---|---|
| Age (years) | 62 (50-71) |
| Gender, M (%n) | 3 (100) |
| BAME (%n) | 2 (66.7) |
| Comorbidities (%n) | |
| Asthma | 0 (0) |
| COPD | 0 (0) |
| Previous pneumothorax | 0 (0) |
| Hypertension | 3 (0) |
| Ischaemic heart disease | 0 (0) |
| Type 2 diabetes mellitus | 0 (0) |
| Smoking history | |
| Current smoker | 0 (0) |
| Ex-smoker (>=10 pack year) | 1 (33.3) |
| Non-smoker or ex-smoker <10 pack year) | 2 (66.7) |
| Severity of acute illness | |
| Hospitalized (n/%) | 3 (100) |
| - Oxygen therapy only (moderate) | 0 (0) |
| - CPAP/ NIV (severe) | 0 (0) |
| - IMV (very severe) | 3 (100) |
| Not hospitalized (moderate) | 0 (0) |
| Length of hospital stay (days) [n=35] | 18 (11-28) |
| Admission bloods [n=3] | |
| White cell count (x10⁹/L) | 9.3 (7.4 – 10.3) |
| Haemaglobin (g/L) | 140 (130 – 151) |
| Platelet (x10⁹/L) | 243 (219 – 524) |
| Neutrophil (x10⁹/L) | 7.7 (6.1 – 9.1) |
| Lymphocyte (x10⁹/L | 0.9 (0.7 – 0.9) |
| D dimer (microgram/L) | 1920 (965 – 2874) |
| Fibrinogen (g/L) | 7.5 (7.2 – 8.6) |
| Ferritin (micrograms/L) | 745 (738 – 6032) |
| Sodium (mmol/L) | 132 (132 – 137) |
| Potassium (mmol/L) | 4.2 (4.2 – 4.9) |
| Urea (mmol/L) | 5 (4.4 – 8.1) |
| Creatinine (micromol/L) | 95 (95 – 135) |
| Egfr (ml/min/1.73m²) | 70 (49 – 81) |
| Albumin (g/dL) | 33 (31 – 34) |
| C-reactive protein (mg/L) | 229 (218 – 255) |
| Treatment during follow up period | |
| Steroids | 1 (33.3) |
| Antibiotics | 1 (33.3) |
| 1 year follow up CT imaging (n=3) | |
| Days from admission to follow up CT | 388 (382 – 389) |
| Days from discharge to follow up CT | 371 (354-377) |
| 1 year bronchoalveolar lavage fluid sampling (n=3) | |
| Days between admission and sampling | 410 (402 – 412) |
| Days between discharge and sampling | 392 (374 – 401) |
| 1 year Lung function tests [n=3] | |
| Days between admission and lung function tests | 394 (389 – 409) |
| Days between discharge and lung function tests [ | 378 (366-391) |
| FEV1 (L) | 3.3 (2.9-3.9) |
| % predicted FEV1 | 104 (97-123) |
| FVC (L) | 3.9 (3.5 – 4.4) |
| % predicted FVC | 101 (91 – 106) |
| TLC (L) | 5.7 (5.6 – 6.7) |
| % predicted TLC | 94 ( 91 – 95) |
| TLCO ml/min/mmHg | 8.8 (7.3 – 9.9) |

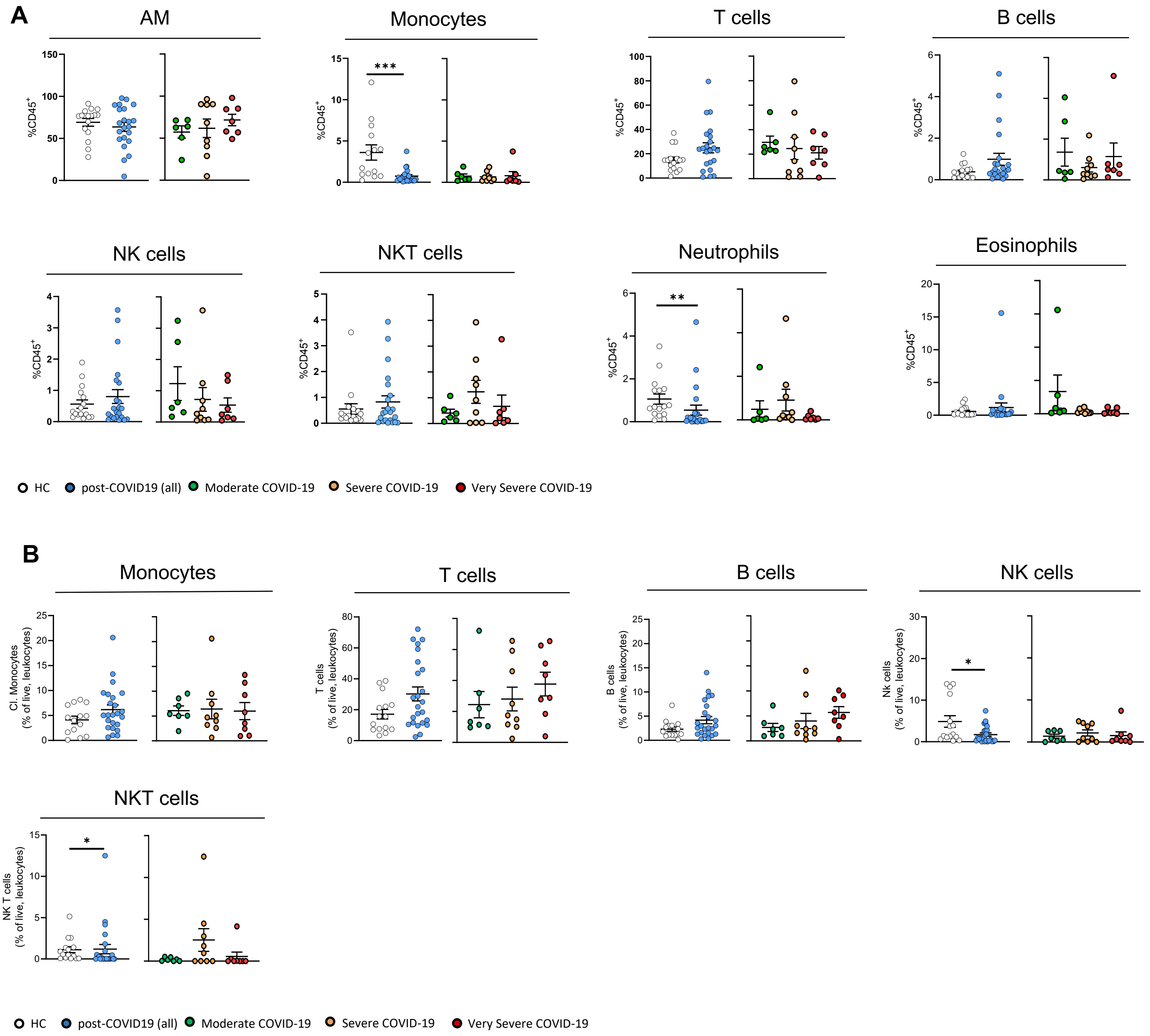

**Figure S1: Flow gating strategy and immune cell proportions in post-COVID19 BAL**

(A) Proportional representation of immune cell populations in BAL from healthy controls and post-COVID19 patients. AM = airway macrophages. (B) Proportions of immune populations (% of live, leukocytes) in peripheral blood from healthy controls and post-COVID19 patients. $CD14^+$ classical monocytes, $CD3^+$ T cells, $CD19^+$ B cells, $CD56^+$ NK cells, $CD3^+/CD56^+$ NKT cells. Post-COVID19 patients were split by severity of acute disease.

Data are presented as mean ± SEM. Healthy controls n = 16, post-COVID-19 patients n = 22, moderate n = 6, severe n = 9, very severe n = 7. Statistical significance was tested by Mann Whitney U test, Kruskal Wallis test + Dunn's multiple comparison test or Spearman Rank test. $*P <$ $**P < 0.01$.

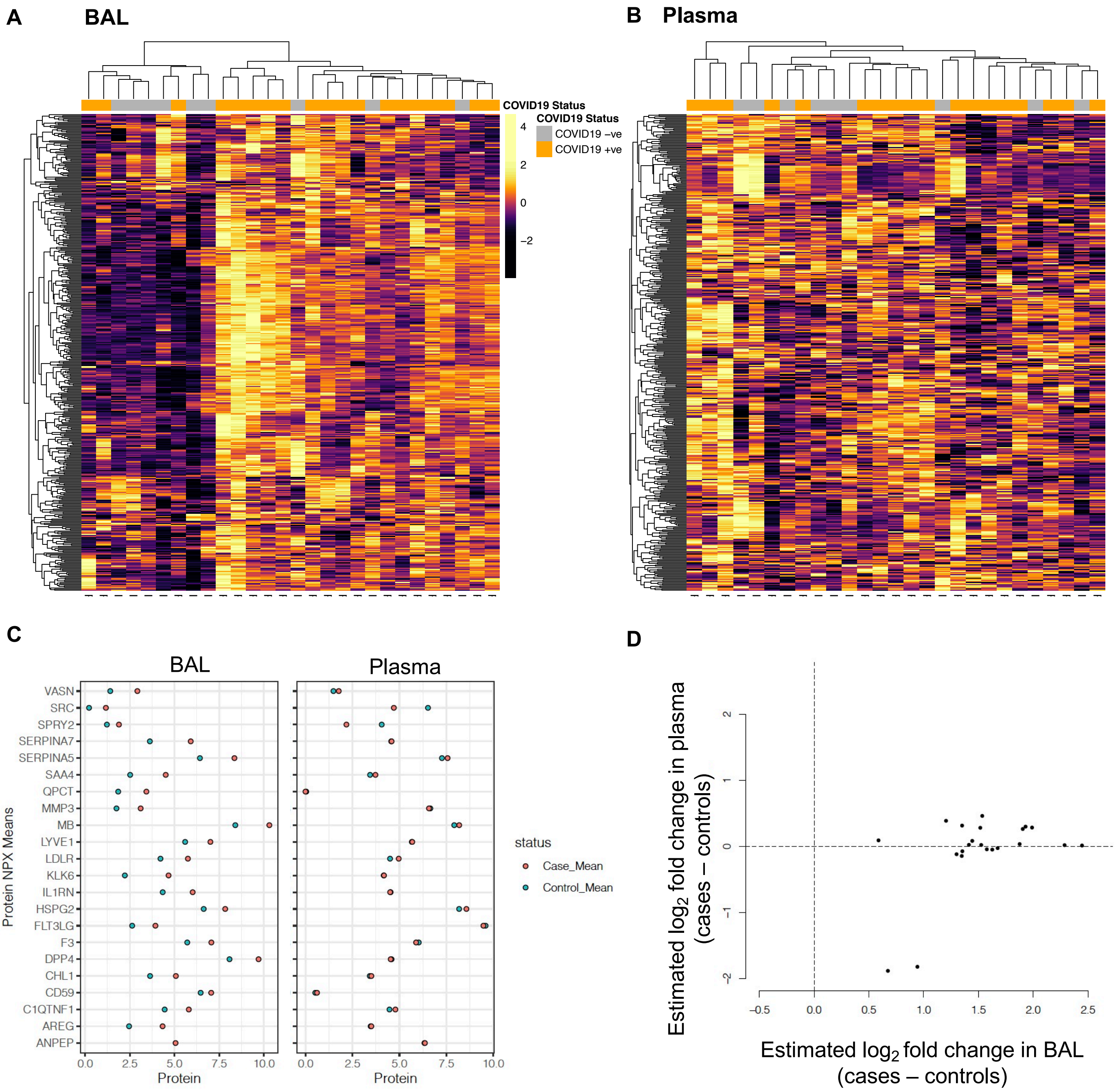

**Figure S2. Post-COVID19 effects on the airway proteome are not mirrored in the blood**
**(A-B)** Hierarchical clustering of samples and proteins using all 435 proteins measured in A) BAL and B) plasma. Z score normalized protein levels are shown. **(C)** Median NPX values for the 22 proteins that were significantly differentially abundant (5% FDR) in BAL from post-COVID-19 patients versus healthy controls. **(D)** Comparison of effect size estimates in BAL versus plasma for the 22 proteins. Each point represents a protein. The x-axis shows the estimated $\log_2$ fold change in BAL while the y-axis shows the estimated $\log_2$ fold change in plasma.

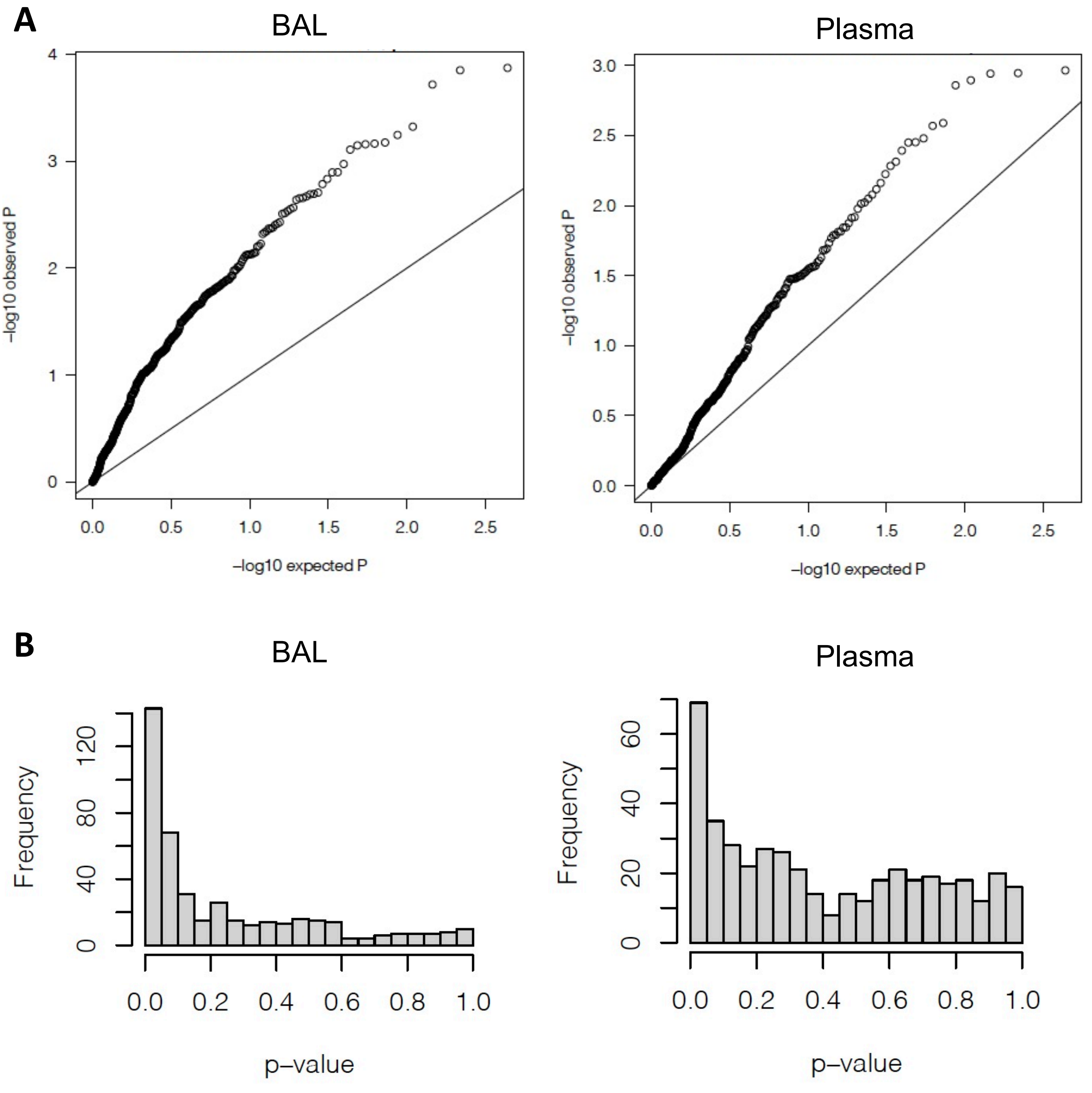

**Figure S3. Statistical evidence for proteomic differences between post-COVID19 and healthy controls**
**(A)** QQ plots showing the expected distribution of –log10 P-values under the null hypothesis of no association between each protein and case/control status compared to the actual –log10 P-values observed in the analysis. The plots shows departure from the diagonal, more marked in BAL. **(B)** Histogram of observed nominal p-values from linear regression of each protein on case/control status. The p-values are not uniformly distributed between 0 and 1. Again this is more marked in BAL.

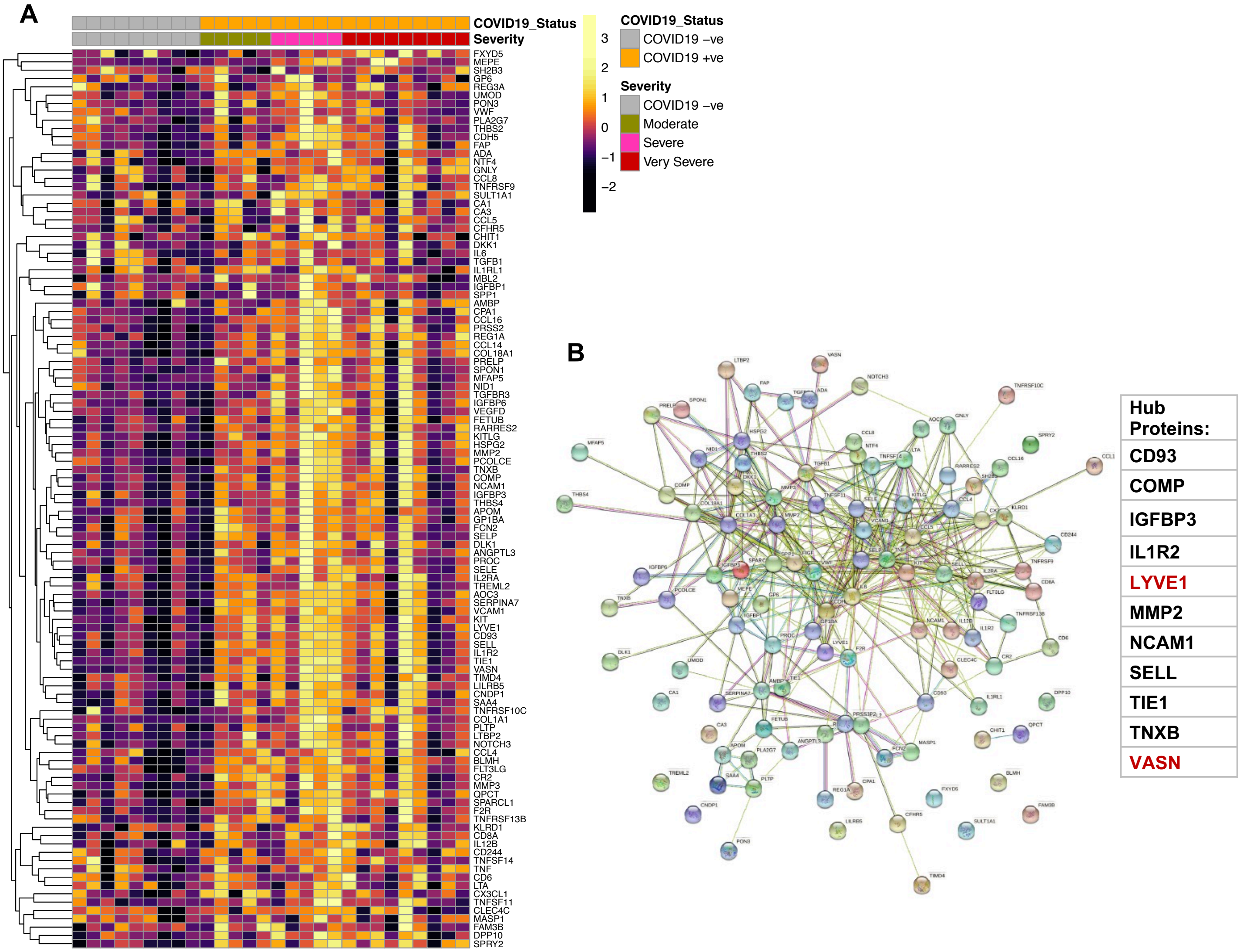

**Figure S4. WGCNA identification of the blue module protein network in the post-COVID19 airway**

**(A)** heatmap displaying Z-score normalised protein abundance for the proteins that form the “blue” eigenprotein module in post-COVID19 and healthy controls in BAL. **(B)** Network representation of proteins in the ‘blue’ module and their interconnections. An edge in the network represents a relationship between proteins defined using String-db. Predicted hub proteins within the network are stated.

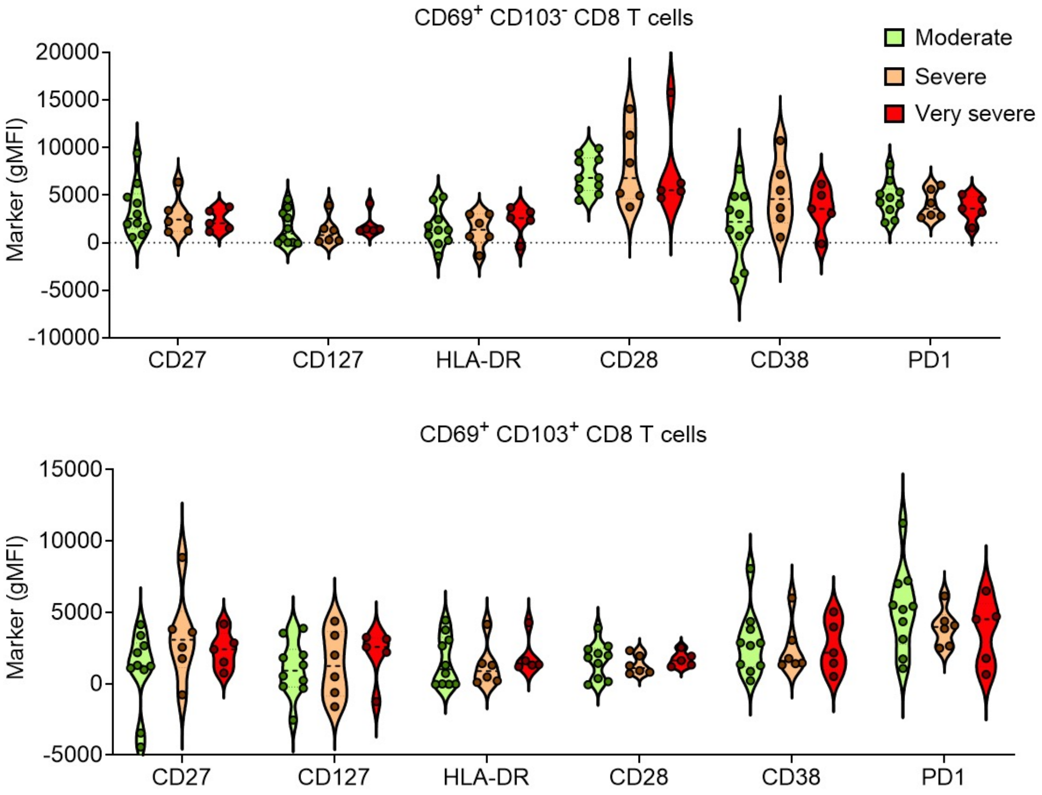

**Figure S5. Activation marker expression on CD8 TRM in the post-COVID19 airway against patient severity**

Airway CD103⁻ (top) and CD103⁺ (bottom) CD69⁺ CD8 T cells were identified in post-COVID19 BAL by spectral deconvolution flow cytometry. Expression of CD27, CD127, HLA-DR, CD28, CD38 and PD-1 was then analysed against acute severity. Geometric mean fluorescence intensity (gMFI) is shown. Each dot represents an individual donor.

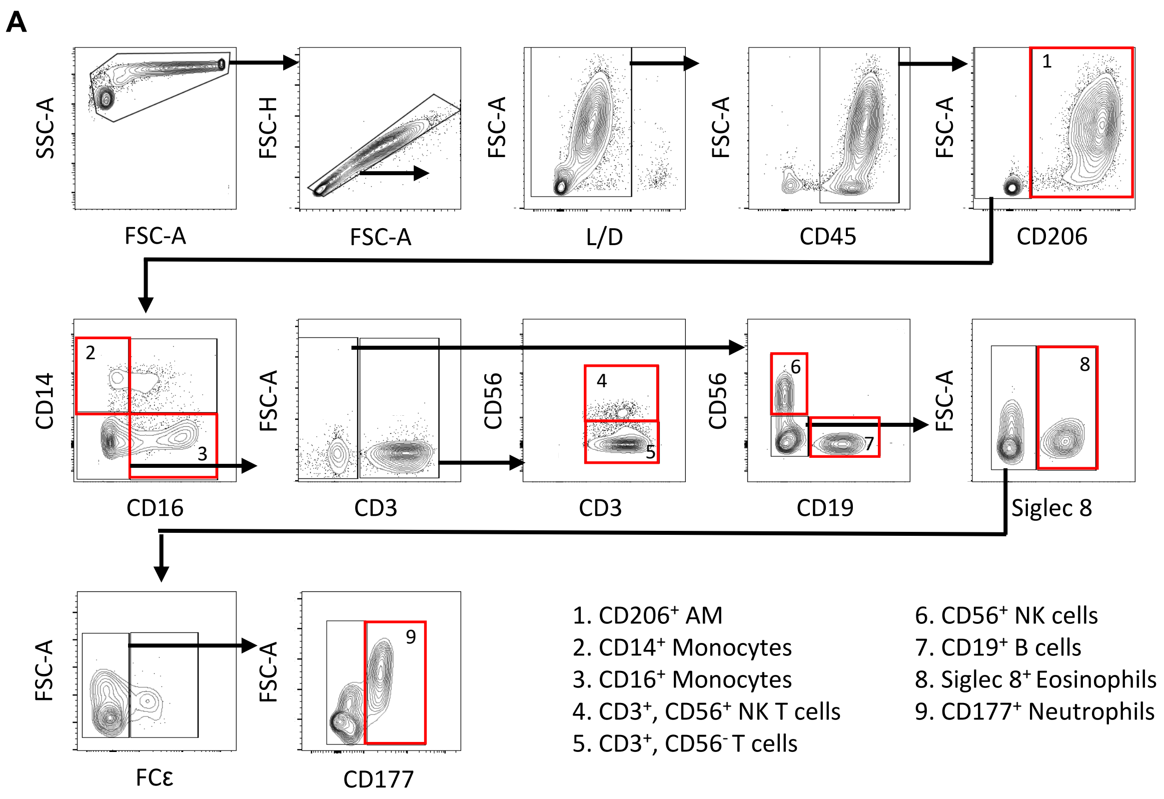

**Methods Figure 1. 11-colour immune phenotyping flow cytometry panel**

Gating strategy to determine immune cell populations in BAL from healthy controls and pCOVID patients. After selecting for live, CD45⁺ cells, the following populations were analysed: CD206⁺ airway macrophages, CD14⁺ classical monocytes, CD16⁺ non classical monocytes, CD3⁺ T cells, CD3⁺/CD56⁺ NK T cells, CD19⁺ B cells, CD56⁺ NK cells, Siglec 8⁺ Eosinophils and CD177⁺ neutrophils.

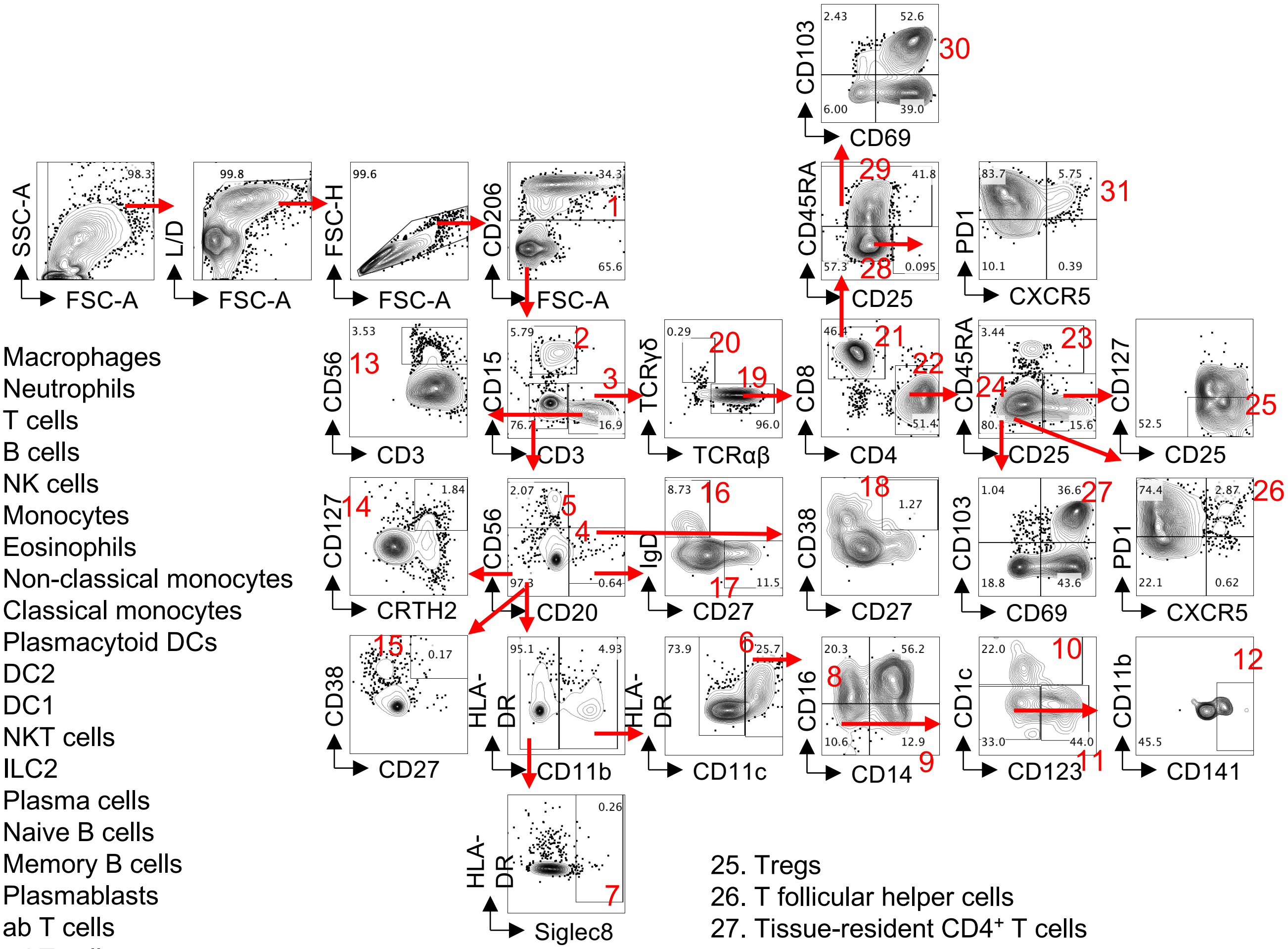

1. Macrophages
2. Neutrophils
3. T cells
4. B cells
5. NK cells
6. Monocytes
7. Eosinophils
8. Non-classical monocytes
9. Classical monocytes
10. Plasmacytoid DCs
11. DC2
12. DC1
13. NKT cells
14. ILC2
15. Plasma cells
16. Naive B cells
17. Memory B cells
18. Plasmablasts
19. ab T cells
20. gd T cells
21. CD8+ T cells
22. CD4+ T cells
23. Naive CD4+ T cells
24. Antigen-experienced CD4+ T cells
25. Tregs
26. T follicular helper cells
27. Tissue-resident CD4+ T cells
28. Antigen-experienced CD8+ T cells
29. Naive CD8+ T cells
30. Tissue-resident CD8+ T cells
31. T follicular cytotoxic cells

**Methods Figure 2. Spectral deconvolution high parameter gating strategy**
Gating strategy for 34-marker spectral deconvolution flow cytometry panel. Post-COVID19 BAL cells are shown.
